# A Hierarchical Visual EEG Framework for the Assessment of Disorders of Consciousness

**DOI:** 10.64898/2026.06.04.26354678

**Authors:** Yuzhen Chen, Qianqian Ge, Hao Li, Xiaoyang Kang, Qi Chen, Weizhong He, Yike Sun, Shangen Zhang, Steven Laureys, Xiaogang Chen, Jianghong He, Xiaorong Gao

**Author notes:** These authors contributed equally to this work (Yuzhen Chen, Qianqian Ge). Correspondence to: Steven Laureys, CERVO Brain Research Centre, Laval University, Québec G1J 2G3, QC, Canada; Coma Science Group, GIGA Consciousness Research Unit, Liège University Hospital, Liège 4000, Belgium; International Consciousness Science Institute, Hangzhou Normal University; Hangzhou 311121, China, Xiaogang Chen, Institute of Biomedical Engineering, Chinese Academy of Medical Sciences and Peking Union Medical College, Tianjin 300192, China, Jianghong He, Department of Neurosurgery, Beijing Tiantan Hospital, Capital Medical University, Beijing 100070, China, Xiaorong Gao, School of Biomedical Engineering, Tsinghua University; Beijing 100084, China.

## Abstract

The objective assessment of patients with disorders of consciousness (DOC) remains a significant clinical challenge. Behavioral scales like the Coma Recovery Scale-Revised (CRS-R) are susceptible to rater subjectivity and have difficulty in detecting patients with cognitive-motor dissociation (CMD), while existing electrophysiological paradigms typically evaluate isolated processing levels, especially in visual functions. To address these limitations, we developed a novel, hierarchical visual EEG framework that evaluates three progressive tiers of visual processing—sensory input, selective attention, and object discrimination—within a single, unified paradigm. This framework uses steady-state and event-related potentials, analyzed with statistical testing and machine learning, to provide objective detection. In a cohort of 85 participants, the framework demonstrated a robust alignment with behavioral CRS-R levels and successfully identified CMD patients missed by bedside behavioral examinations. Notably, model predictions derived from this framework showed a significant correlation with 3-month clinical outcomes. This prognostic utility generalized effectively and remained consistent across distinct EEG acquisition systems in an independent validation cohort of 17 patients. In summary, this work offers electrophysiological validation for the hierarchical design of the CRS-R and provides a practical tool for bedside objective assessment of DOC.

## Introduction

Disorders of consciousness (DOC) refer to a pathological state characterized by an impaired awareness of self or the environment, along with a diminished capacity to respond to external stimuli^1^. Rough estimates suggest that hundreds of thousands of new DOC cases may emerge globally each year, with lifetime care costs for chronic patients potentially exceeding one million US dollars^2–4^, imposing a substantial burden on patients, families, and society. Clinical diagnosis of DOC primarily relies on behavioral scales, with the Coma Recovery Scale–Revised (CRS-R)^5^ being the most widely used. The CRS-R comprises six subscales (auditory, visual, motor, oromotor/verbal, communication, and arousal), with items within each subscale arranged hierarchically. The lowest item in each subscale represents reflexive activity, whereas the highest indicates cognitively mediated behavior^5^. Furthermore, based on the consciousness state and responsiveness, DOC patients are categorized into unresponsive wakefulness syndrome (UWS; preservation of sleep-wake cycles^6^), minimally conscious state (MCS; demonstrating minimal but definite behavioral evidence of self or environmental awarenes ^7^, further subdivided into MCS+ and MCS-based on the presence of command-following behaviors), and emergence from MCS (eMCS; recovery of functional communication or object use).

Compared to behavioral scales, which are susceptible to rater subjectivity, electroencephalography (EEG) provides an objective means to assess consciousness without relying on motor function, and is thus widely utilized in clinical settings^8–11^. It is particularly valuable for detecting patients with cognitive motor dissociation (CMD), a condition defined by the dissociation between preserved cognitive capacity (e.g., command-following) and the lack of purposeful motor response^12–14^. Mirroring the hierarchical design of behavioral scales, research on evoked potentials in EEG has also employed graded paradigms, providing a window into patients’ low-level to high-level sensory and cognitive functions while potentially reducing assessment time. However, existing hierarchical paradigms for DOC assessment have been largely confined to the auditory^15–21^ and somatosensory^22^ modalities, whereas visual paradigms remain predominantly single-level. While recent studies on brain-computer interface (BCI) based on visual evoked potential (VEP) have demonstrated the feasibility of detecting residual consciousness via the visual channel^13,23–34^, these works have predominantly focused on engineering-oriented reports. They lacks the capacity for a systematic assessment of multi-level visual functions within a unified paradigm, as well as correlation with the hierarchical levels of clinical behavioral scales.

To address this gap, we propose a hierarchical visual electrophysiological assessment framework. The contributions of this framework are fourfold: 1) Multi-level assessment: The use of a unified visual paradigm to simultaneously probe three hierarchical levels of processing—from basic sensory input to selective attention and visual discrimination; 2) Clinical mapping: Direct mapping of electrophysiological features to CRS-R behavioral levels, providing electrophysiological validation for the scale’s hierarchical design; 3) CMD detection: Objective identification of CMD patients missed by standard behavioral assessment; 4) Prognostic relevance: Establishment of links between electrophysiological features and patient clinical outcomes.

Specifically, within this framework, we designed a novel hierarchical visual paradigm. Novel stimuli are embedded within the two rapid serial visual presentation (RSVP) targets, thereby incorporating three stimulus elements corresponding to sensory input, selective attention, and object discrimination in visual processing. For the distinct electrophysiological responses evoked by these different hierarchical elements, we further developed a discrimination method based on statistical testing and machine learning. This method is used to determine whether an individual possesses specific levels of visual function, thereby inferring their state of consciousness. Our framework was first validated in a cohort of 85 participants, followed by cross-device generalizability testing in an additional 17 participants. The results demonstrated that the levels of visual function identified by this framework show high concordance with behavioral scales and are significantly correlated with the clinical diagnosis and prognosis. The hierarchical electrophysiological framework proposed here combines interpretability with potential for generalization, showing promise for development into an objective and efficient new tool for bedside auxiliary diagnosis and prognosis assessment in DOC.

## Materials and methods

### Participants

Participants were recruited from Beijing Tiantan Hospital and Beijing Tieying Hospital between January 2024 and October 2025. Inclusion criteria for DOC patients were: 1) age between 14 and 65 years; 2) diagnosis of a DOC after emerging from coma; 3) time post-onset of more than one month; and 4) acute-onset DOC (e.g., due to traumatic brain injury, stroke, or hypoxic-ischemic encephalopathy) rather than disorders caused by progressive neurodegenerative diseases. Exclusion criteria included inability to maintain an eye-open state, a history of skull defect or cranioplasty that could significantly affect EEG signal quality, or other conditions that would prevent compliance with the testing procedure. This study was reviewed and approved by the Medical Ethics Committee of Beijing Tiantan Hospital, Capital Medical University, Beijing, China (Approval No. 2025-357-02). All procedures were conducted in accordance with the relevant guidelines and regulations, and informed consent was obtained from all participants or their legal surrogates.

A total of 102 participants were recruited. Data from 85 participants were acquired using a saline-based EEG acquisition system (HydroCel Geodesic Sensor Net, EGI, Eugene, USA). This cohort included 69 patients with DOC (43.30 ± 14.45 years, 50 males), one patient with patient with locked-in syndrome (aged 50–55, male), and 15 healthy controls (HC; 30.07 ± 7.25 years, 13 males). Data from the remaining 17 DOC patients (45.41 ± 9.86 years, 10 males) were acquired using a conductive paste-based EEG system (SynAmps2, Neuroscan, El Paso, USA). The distribution of diagnostic labels and visual scores for all participants is shown in Table 1. Detailed clinical information is provided in Supplementary Table 1. Three months after the experiment^13^, among the 62 DOC patients (excluding eMCS) from the EGI cohort, 9 had a favorable prognosis (indicated by an improvement in consciousness diagnosis, e.g., from UWS to MCS-), 39 had a poor prognosis, and 14 were lost to follow-up. Among the 17 patients assessed with the Neuroscan system who underwent deep brain stimulation treatment after admission, 8 had a favorable prognosis, 8 had a poor prognosis, and 1 was lost to follow-up.

### Visual Function Assessment in CRS-R

For the assessment of visual function^5^, the examiner rapidly brings a finger or object from the side towards the patient’s eye to present a threat and observes for eyelid flutter or blinking (score 1). Subsequently, a small mirror or a video potentially of interest to the patient is presented 30-40 cm in front of the patient’s face and moved quickly in different directions. The examiner observes whether the patient can re-fixate from the initial point of regard to the new target location and sustain fixation for at least 2 seconds (score 2). Further, the object is moved slowly and smoothly along the midline, and the examiner observes whether the patient’s eyes can follow the movement smoothly and continuously over 45° in the horizontal or vertical plane (score 3). Next, an object potentially of interest is presented within the patient’s visual field, and a verbal command or gesture is given for the patient to reach for/touch the object. The examiner observes whether the patient’s limb moves towards the object (score 4). Finally, two functionally different everyday objects are placed in distinct locations. A verbal command is given for the patient to look at or touch the different objects, and the examiner observes for any reproducible and correct behavior (score 5).

### Apparatus

This study employed EGI and NeuroScan systems for EEG data acquisition. The EGI system was used in either a 128-channel or 256-channel configuration (Supplementary Fig. 1A and B), with a sampling rate of 250 Hz. The reference electrode was positioned at Cz, and the ground electrode was placed at the midpoint between CPz and Pz. The NeuroScan system was used in a 64-channel configuration (Supplementary Fig. 1C), with a sampling rate of 1 kHz, the reference electrode placed on the vertex, and the ground electrode positioned at the midpoint between FPz and Fz. Visual stimuli for this study were programmed in MATLAB and presented on a 16-inch monitor (ThinkVision M15, Lenovo, Beijing, China), which was either mounted on a stand or held by hand, positioned 40-60 cm in front of the participant.

### Paradigm

In the paradigm used in this study (Fig. 1A and B), two targets, each 800 × 800 pixels in size, were presented at the one-quarter and three-quarter points along the long edge of the display screen. Both targets were picture sequences with a fixed frequency in an “On-Off” mode. The presentation frequency for the left target was f1 = 6 Hz, and for the right target was f2 = 7.6 Hz. The selection of these two frequencies was based on experimental settings from a previous study^35^. The paradigm consisted of 40 blocks. Each block contained two stimulation trials. Each trial comprised 1 s of instruction, 5 s of stimulation, and 2 s of rest. In the first trial of a block, one of the two targets, selected at random, displayed a sequence of randomly selected colored abstract pictures. Within the 2-4 s time window of this sequence, one randomly chosen abstract picture was replaced with a random human face picture. The other target in this trial displayed only a sequence of black-and-white pictures. In the second trial of the same block, the positions of the colored and black-and-white picture sequences were swapped. The color images used in this paradigm were generated by Midjourney (https://www.midjourney.com/), including 50 abstract images (Supplementary Fig. 6A) and 3 images (Supplementary Fig. 6B) each for three age groups (children, middle-aged, and elderly) and two genders (male and female).

**Figure 1.**
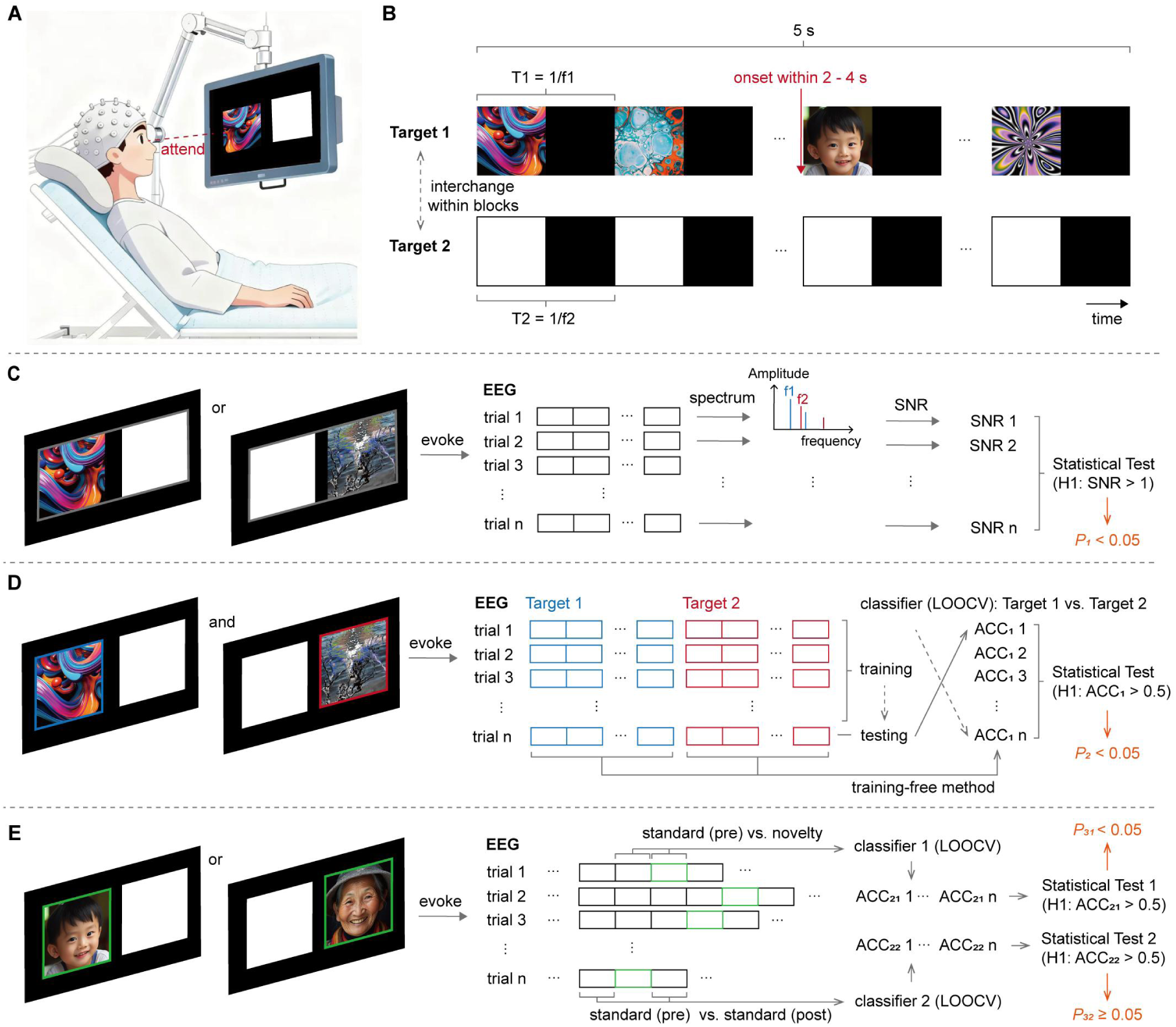
Experimental paradigm and the corresponding evoked potential discriminant framework. (**A**) Schematic of stimulus presentation. Participants were instructed to attend to the colored target on the screen. (**B**) Schematic of the paradigm flow. A sequence of colored abstract pictures (with one picture replaced by a human face) and a sequence of black-and-white pictures are presented on the two targets in random order. Facial images shown are computer-generated (via *Midjourney*) and are not photographs of real people. (**C**) Picture sequences presented on the screen at fixed frequencies elicit a SSVEP containing the frequency information. The presence of the SSVEP is detected by calculating the SNR at the specific frequency and determining if it is significantly greater than 1. (**D**) The left and right targets on the screen are tagged with different frequencies. A classifier is constructed to discriminate between neural responses when the colored pictures are on the left versus on the right. The participant’s ability to attend to the colored pictures as instructed is assessed by determining if the classifier’s accuracy (ACC_1_) is significantly greater than 0.5 (chance level for a binary task). (**E**) The human face picture embedded among the colored pictures serves as a novel stimulus, eliciting an ERP distinct from the response to standard stimuli. The accuracy (ACC_21_) of a classifier discriminating responses to the novel versus a standard stimulus is used to detect a change in response after face onset, determined by testing if ACC_21_ is significantly greater than the chance level for a binary task (0.5). The accuracy (ACC_22_) of a classifier discriminating responses to two standard stimulus segments (one before and one after the novel stimulus) serves as a control, and its non-significance helps rule out response changes caused by non-cognitive factors (e.g., eye closure or gaze shift), thereby confirming the specificity of the ACC_21_ result.

Before the experiment, medical staff instructed participants via on-screen text and verbal commands to attend to the target containing the colored pictures and to ignore the target with black-and-white pictures during the experiment. The use of colored versus black-and-white pictures was intended to attract the attention^36^ of participants who might have difficulty seeing clearly, hearing clearly, or understanding the task requirements, thereby achieving the goal of overt or covert attention. For DOC patients, medical staff confirmed they were in an awake state before the experiment and would rouse any patient showing signs of falling asleep (eye closure) during the procedure.

### Data Pre-processing

The acquired EEG data were notch-filtered at 50 Hz and band-pass filtered between 1 and 50 Hz. Data from different acquisition systems were resampled to a unified rate of 250 Hz. Subsequently, bad channels were identified and reconstructed using spherical interpolation via the EEGLAB toolbox, and all data were re-referenced to the average of the left and right mastoids. To facilitate unified analysis, channels that were common across different acquisition configurations and conformed to the naming conventions of the 10-20 system were extracted for subsequent processing (Supplementary Fig. 1).

### Evoked Potential Identification

The paradigm incorporates three stimulus elements corresponding to the three hierarchical levels of visual function: sensory input, selective attention, and visual discrimination. The evoked potentials associated with each level possess distinct characteristics and require different methods for detection (see Supplementary Materials for details).

#### 1) Sensory Input

The first level (Fig. 1C) pertains to basic visual sensory input and assesses the primary processing capacity of the visual system for incoming light signals. When picture sequences are presented on the screen at a fixed frequency, they drive neurons in the primary visual cortex to generate frequency-following activity, thereby producing SSVEP phase-locked to the stimulation frequency in the EEG signal^37,38^. The strength of the neural entrainment to the driving frequencies was quantified by calculating the signal-to-noise ratio (SNR)^39,40^ of the SSVEP using canonical correlation analysis (CCA)^41^. For each participant, we statistically tested whether the SNR was significantly greater than 1 (the null response level) to determine the presence of a basic sensory response^42^. The resulting p-value (*P_1_*) from this test provided a binary outcome for the presence or absence of significant sensory input function.

#### 2) Selective Attention

The second level (Fig. 1D) probes visual selective attention. This is a core cognitive control function that enables the brain to allocate processing resources to task-relevant stimuli while suppressing distractions^43^. When attention is directed to one of the two frequency-tagged targets, top-down modulation enhances the neural response at the attended location^44^. Our framework infers the attended location by constructing a classifier to decode the SSVEP signal. To ensure methodological robustness and minimize algorithm bias, we employed three established algorithms: Filter Bank CCA (FBCCA, a training-free method)^45^, Task-Related Component Analysis (TRCA)^46,47^, and Task-Discriminant Component Analysis (TDCA)^48^ (training-based methods). The classification accuracy (ACC_1_) reflects the ability to modulate attention spatially. This accuracy was statistically compared against the chance level (0.5 for a binary task) to obtain a p-value (*P_2_*). A significant ACC_1_ (*P_2_* < 0.05) indicates the presence of covert or overt spatial attention, marking a transition from passive perception to active selection.

#### 3) Visual Discrimination

The third level (Fig. 1E) assesses the visual system’s ability to discriminate and categorize complex stimuli, a process often linked to higher-order cognitive processing. Our framework introduces a human face as a novel stimulus within the standard stimulus stream. Detection of this deviant stimulation by the brain elicits a specific ERP^49–51^. To objectively quantify this response, we implemented a dual-classifier approach with statistical controls. First, a classifier (based on TRCA) was built to discriminate between the neural response epochs following the novel stimulus and those following a standard stimulus, yielding accuracy ACC_21_. Second, a control classifier was built to discriminate between responses to two standard stimulus segments (one before and one after the novel stimulus), yielding accuracy ACC_22_. The purpose of ACC_22_ is to rule out non-specific neural changes (such as eye closure or gaze shifts) that could lead to false-positive significance in ACC_21_. Therefore, a significant ACC_21_ (tested against chance level, yielding *P_31_*) coupled with a non-significant ACC_22_ (yielding *P_32_*) specifically indicates that the brain not only registered but could discriminate the novel stimulus, signifying preserved visual discrimination ability.

### Statistical Analysis

#### Feature Analysis and Group Comparisons

Participants recruited in this study were grouped according to their diagnostic labels and visual scores, and differences in metrics between groups were compared. The sole patient with a visual score of 2 was pooled with the score-1 group to facilitate group-wise analysis. For comparisons among three or more groups, one-way ANOVA with post-hoc tests (Bonferroni correction) was employed. For direct comparisons between two groups, an independent samples t-test was used. Additionally, the number of participants with significant versus non-significant metrics at each level within each group was counted, and the chi-squared test was applied to examine the significance of these distributions.

For characterizing the ERP evoked by the novel stimulus, correlation analysis was used to examine the correlation between consecutive SSVEP cycles. Furthermore, for data with the SSVEP component removed, a permutation test method analogous to temporal clustering^52^ was employed for ERP detection. First, at the single-trial level, a t-statistic was computed for each time point. All contiguous time points where the t-statistic exceeded the confidence threshold (α = 0.05) were defined as a “cluster”, and the sum of the t-statistics within this cluster was calculated as the cluster mass. Subsequently, each trial was randomly assigned a coefficient of either +1 or -1. Multiplying the data by these coefficients generated a set of permuted data, and the maximum cluster mass within this permuted data was recorded. This permutation procedure was repeated 1000 times to obtain a null distribution of the maximum cluster mass. The cluster mass of the actual observed data was then compared against this distribution. If the mass of an actual cluster exceeded 95% of the values in the permutation distribution (corresponding to a confidence level of 0.05), that cluster was considered significant, indicating the presence of a significant non-zero ERP component during that time period.

The extracted features were categorized into two types: numerical indexes (SNR, ACC_1_, ACC_21_, ACC_22_) and binary features (*B*(*P_1_*), *B*(*P_2_*), *B*(*P_31_*, *P_32_*)). Here, *B*(*P*) denotes the binarization of p-values based on the criteria described earlier (where α represents the confidence level, and *NA* indicates a missing value):

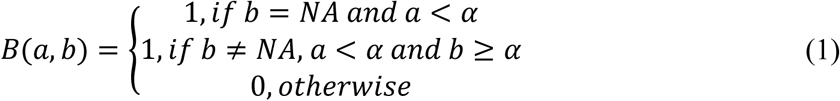

To analyze the association between evoked potential features and clinical information, linear regression analysis was performed between the features and the continuous visual scores. Furthermore, correlation analysis was conducted between the age and the onset duration of DOC patients, and the evoked potential features. Patients were also grouped by etiology, and one-way ANOVA was used to compare features across different etiology groups. Features were extracted using different numbers of channels, and the influence of channel count was assessed using ANOVA with post-hoc tests (Bonferroni correction).

#### Classification Model and Prognostic Analysis

To investigate the efficacy of our framework in predicting the consciousness state, participants were grouped to form data belonging to three categories: HC, MCS (a combination of MCS- and MCS+), and UWS. Two classification models were constructed based on the numerical features and the binary features, respectively.

For the numerical features, a model was constructed using a decision tree algorithm. The model was trained and evaluated on the EGI data employing a 5-fold cross-validation strategy; this process involves partitioning the data into five subsets, each serving in turn as the validation set while the remaining four form the training set, to obtain a robust estimate of classification accuracy. Subsequently, a hard voting ensemble strategy was employed to enhance the model’s generalizability and robustness. In this strategy, the base model (trained on the EGI data) generates predictions, and the final classification for each sample in the Neuroscan dataset is determined by the majority vote across all base predictors, thereby applying the knowledge learned from EGI to the Neuroscan data. The corresponding classification accuracy on the Neuroscan data was then computed.

For the binary features, a model classification criterion was established based on a priori knowledge: the binary indicators from the three levels were summed. A participant with a sum of 3 was classified as HC; a sum of 2 as MCS; and a sum less than 2 as UWS. The binary feature model was also applied separately to the EGI data and the Neuroscan data, and the corresponding accuracy was calculated.

Finally, we tallied the number of individuals with good versus poor prognosis within the group predicted by the models to have a higher consciousness state (e.g., UWS predicted as MCS) and within the group predicted to have the same or a lower consciousness state. The chi-squared test was employed to examine the significance of the difference in these counts, thereby evaluating the models’ potential for prognosis prediction.

## Results

### Level 1: Sensory Input

We used the SNR of the SSVEP to quantify basic visual sensory input function. Based on individual SNR data (Supplementary Fig. 2A), we analyzed the group-averaged SNR spectrum (Fig. 2A and B; statistical test results are in Supplementary Table 2). Peaks corresponding to the stimulation frequencies (f1, f2) and their harmonics (integer multiples of f1 and f2) are visible in the group SNR spectrum, generally showing a trend of decreasing response strength with increasing harmonic order. Furthermore, it can be roughly observed that patients with lower diagnostic levels or visual scores exhibited weaker SSVEP responses. Conversely, groups with higher levels showed significant SSVEP responses, with spectral differences due to attention directed to different target sides.

**Figure 2.**
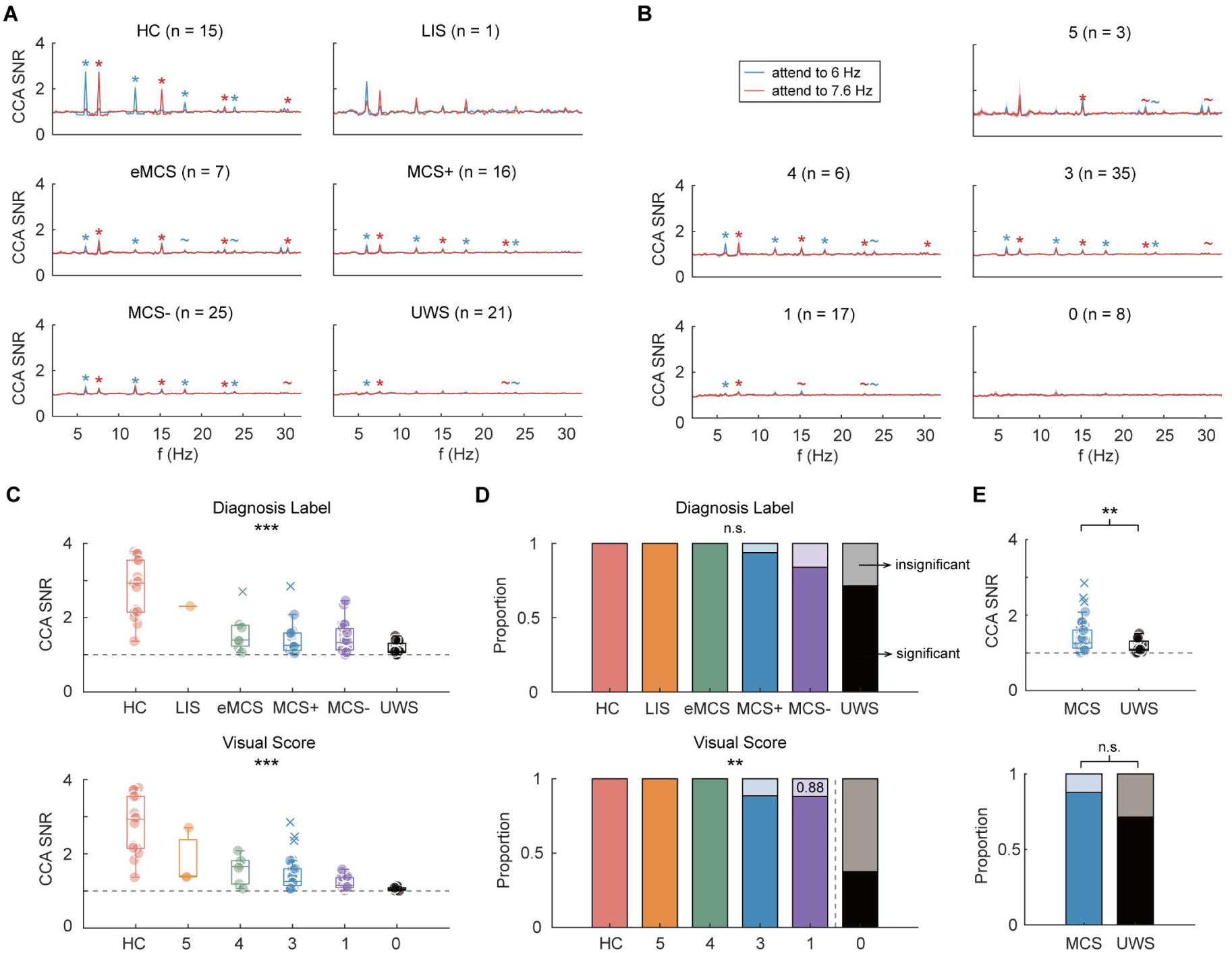
SNR results of the SSVEP across different groups. Group-averaged SNR spectra for (**A**) different diagnosis label groups and (**B**) different visual score groups. Shaded areas represent the standard error of the mean. (**C**) SNR corresponding to the stimulation frequency across different groups. Results of multiple comparisons (Bonferroni corrected) are shown in table S3. (**D**) The proportion of participants with significant SNR in each group. Dark and light colored areas represent the proportion of participants with and without significant SNR, respectively. The grey dashed line in the lower panel demarcates groups with and without a visual startle reflex according to the CRS-R. (**E**) SNR values and the proportion of participants with significant SNR for MCS and UWS groups. For clarity of annotation in panel (**A**), responses with *P* < 0.05, *P* < 0.01, and *P* < 0.001 are all indicated by an asterisk (*). In panels (**C**-**E**): not significant (n.s.), *P* > 0.1; ∼, *P* < 0.1; *, *P* < 0.05; **, *P* < 0.01; ***, *P <* 0.001.

The maximum SNR corresponding to the fundamental stimulation frequency within the SSVEP was extracted for characterization. Figure 2C shows significant differences in SNR among groups with different diagnosis labels [*F*(5,79) = 23.572, *P* < 0.001, *η*^2^ = 0.598; one-way ANOVA] and visual score groups [*F*(5,78) = 24.468, *P* < 0.001, *η*^2^ = 0.612; one-way ANOVA]. However, post-hoc tests (Bonferroni correction) revealed that this difference primarily stemmed from comparisons between the HC group and other groups (Supplementary Table 3). We compared the SNR to the null response level (SNR = 1) and calculated the corresponding p-value (*P_1_* in Fig. 1C). The number of participants with SNR significantly greater than 1 (*P_1_* < 0.05) and those without (*P_1_* ≥ 0.05) was counted and compared across groups. Figure 2D showed that the number of participants with significant SNR gradually decreased as the diagnosis level or visual score declined. Regarding diagnosis labels, the vast majority of participants at the UWS level and above had significant SNR. For visual scores, the number of participants with significant SNR differed significantly among groups (χ² = 20.886, *P* = 0.001; Chi-squared test). Notably, in groups with scores greater than 0 (indicating the presence of a visual startle reflex), over 88% of participants had significant SNR. This demonstrates a high consistency between using SSVEP SNR and the clinical CRS-R visual subscale in assessing basic visual sensory input function. Additionally, a subset of the group with a score of 0 was able to produce a basic response to visual input, suggesting that SSVEP SNR might be more sensitive than startle reflex examination in the CRS-R when assessing sensory input function.

Furthermore, to characterize the potential of SNR in indicating the consciousness state, we merged MCS- and MCS+ into a single MCS group and compared their SNR with the UWS group (Fig. 2E). The results showed a significant difference in SNR between MCS and UWS (*t* = 2.808, *P* = 0.007, *Cohen’s d* = 0.754; two-tailed unpaired t-test). The proportion of participants with significant SNR was also higher in the MCS group than in the UWS group, although this difference was not statistically significant (χ² = 2.552, *P* = 0.110; Chi-squared test). These results indicate that SSVEP SNR has a certain, albeit potentially limited, utility in indicating the consciousness state.

### Level 2: Selective Attention

For attending to the color targets, we processed the data from each participant using the algorithms described above (Supplementary Fig. 2B) and calculated the group-level performance. The results indicated a general trend where the longer the attention was maintained on the stimulus, the higher the accuracy rate across all groups (Fig. 3A and B; Supplementary Table 4). At the group-average level, the training-based algorithms yielded higher accuracy than the training-free algorithm in most cases. This suggests that the evoked potential patterns in a subset of DOC patients also exhibit consistency in fixation behavior across trials, and demonstrate differences under two attention tasks.

**Figure 3.**
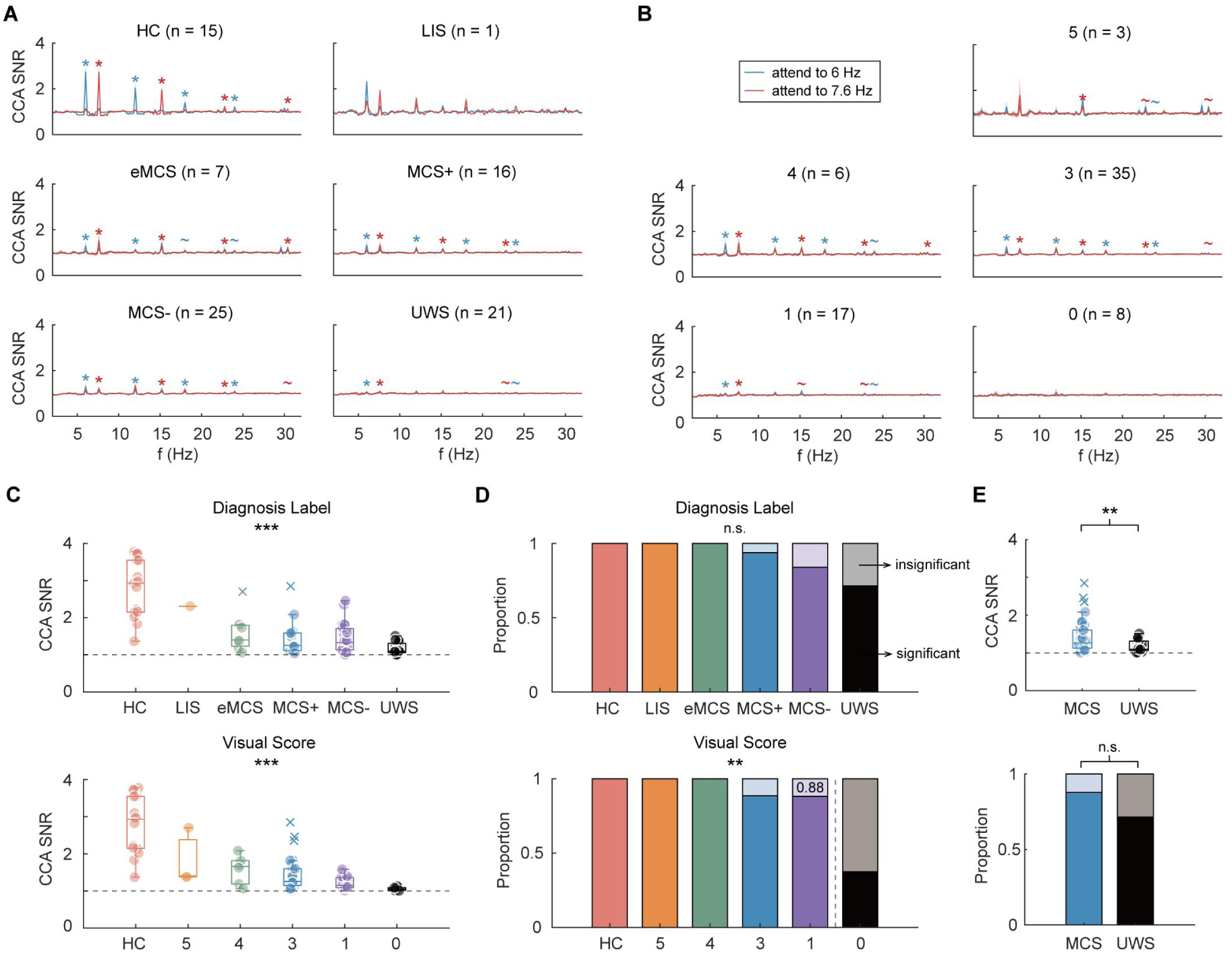
ACC_1_ of the classifier discriminating left versus right target stimulation across groups. Accuracy-time curves for three algorithms in (**A**) different diagnosis label groups and (**B**) different visual score groups. Shaded areas represent the standard error of the mean. (**C**) ACC_1_ values across different groups. Results of multiple comparisons (Bonferroni corrected) are shown in table S5. (**D**) The proportion of participants with significant ACC_1_ in each group. Dark and light colored areas represent the proportion of participants with and without significant ACC_1_, respectively. The grey dashed line in the lower panel demarcates groups with and without object tracking behavior according to the CRS-R. (**E**) ACC_1_ values and the proportion of participants with significant ACC_1_ for MCS and UWS groups. For clarity of annotation in panel (**A**), responses with *P* < 0.05, *P* < 0.01, and *P* < 0.001 are all indicated by a solid circle (•). In panels (**C-E**): not significant (n.s.), *P* > 0.1; ∼, *P* < 0.1; *, *P* < 0.05; **, *P* < 0.01; ***, *P <* 0.001.

The best average accuracy from the final second of data across the three algorithms was extracted for analysis. We found that ACC_1_ differed significantly among groups with different diagnostic levels [*F*(5, 79) = 22.361, *P* < 0.001, *η*^2^ = 0.586; one-way ANOVA] and visual scores [*F*(5, 78) = 23.286, *P* < 0.001, *η*^2^ = 0.600; one-way ANOVA] (Fig. 3C and Supplementary Table 5). The proportion of participants with *P_2_* below the significance threshold (α = 0.05) was calculated for each group. This proportion exhibited a stepwise pattern (Fig. 3D) and showed significant differences across groups defined by both diagnosis labels (χ^2^ = 25.559, *P* < 0.001; Chi-squared test) and visual scores (χ^2^ = 26.040, *P* < 0.001; Chi-squared test). Specifically, the vast majority of individuals with a diagnosis above UWS and with a visual score greater than 2 showed significant ACC_1_. This indicates that the selective attention function evaluated by our framework is common in groups with better-preserved function and aligns closely with the visual tracking function assessed by the CRS-R. Concurrently, a small subset of UWS patients and those with a visual score of 0 also demonstrated significant ACC_1_, suggesting they retained the capacity for two distinct attentional behaviors. This implies our framework holds potential for detecting covert attention or CMD. Furthermore, a direct comparison between MCS and UWS groups (Fig. 3E) revealed a significant difference in ACC_1_ (*t* = 3.165, *P* = 0.002, *Cohen’s d* = 0.849; two-tailed unpaired t-test), with an effect size larger than that observed for SNR. The difference in the proportion of participants with significant ACC_1_ between these groups was also significant (χ² = 12.769, *P* < 0.001; Chi-squared test). These results demonstrate that ACC_1_ possesses strong potential for discriminating levels of consciousness.

### Level 3: Visual Discrimination

To elicit an ERP response associated with visual discrimination, we embedded a single concrete human face picture (novel stimulus) within the sequence of colored abstract pictures (standard stimulus) that participants were instructed to attend to. To confirm the presence of this ERP component, we first analyzed data from the HC. As the presentation of the novel stimulus did not alter the picture presentation frequency, the elicited ERP is embedded within the ongoing SSVEP (blue solid line in Fig. 4A). The SSVEP is a steady-state response with high signal similarity across its cycles. The data were segmented according to the period corresponding to the stimulation frequency, and the correlation between consecutive segments was calculated sequentially. Following the appearance of the novel stimulus, this correlation level dropped sharply from a high level to a lower one and recovered after several hundred milliseconds (red dashed line in Fig. 4A). A matrix constructed from the correlation coefficients between all data segments also showed that segments before the novel stimulus were highly correlated with each other, whereas segments after the novel stimulus exhibited reduced correlation with the rest of the data (Fig. 4C; the average results of all channels are shown in Supplementary Fig. 3D). Subtracting the data following a standard stimulus from the data following the novel stimulus allowed for further isolation of the ERP component: activity at the representative frontal channel (Fz) showed significant fluctuations between 200-600 ms, and activity at the typical occipital channel (Oz) persisted up to 800 ms (Fig. 4B; results for all channels are shown in Supplementary Fig. 3A). Topographic maps also revealed neural activity in the frontal, parietal, occipital, and temporal regions (Fig. 4D).

**Figure 4.**
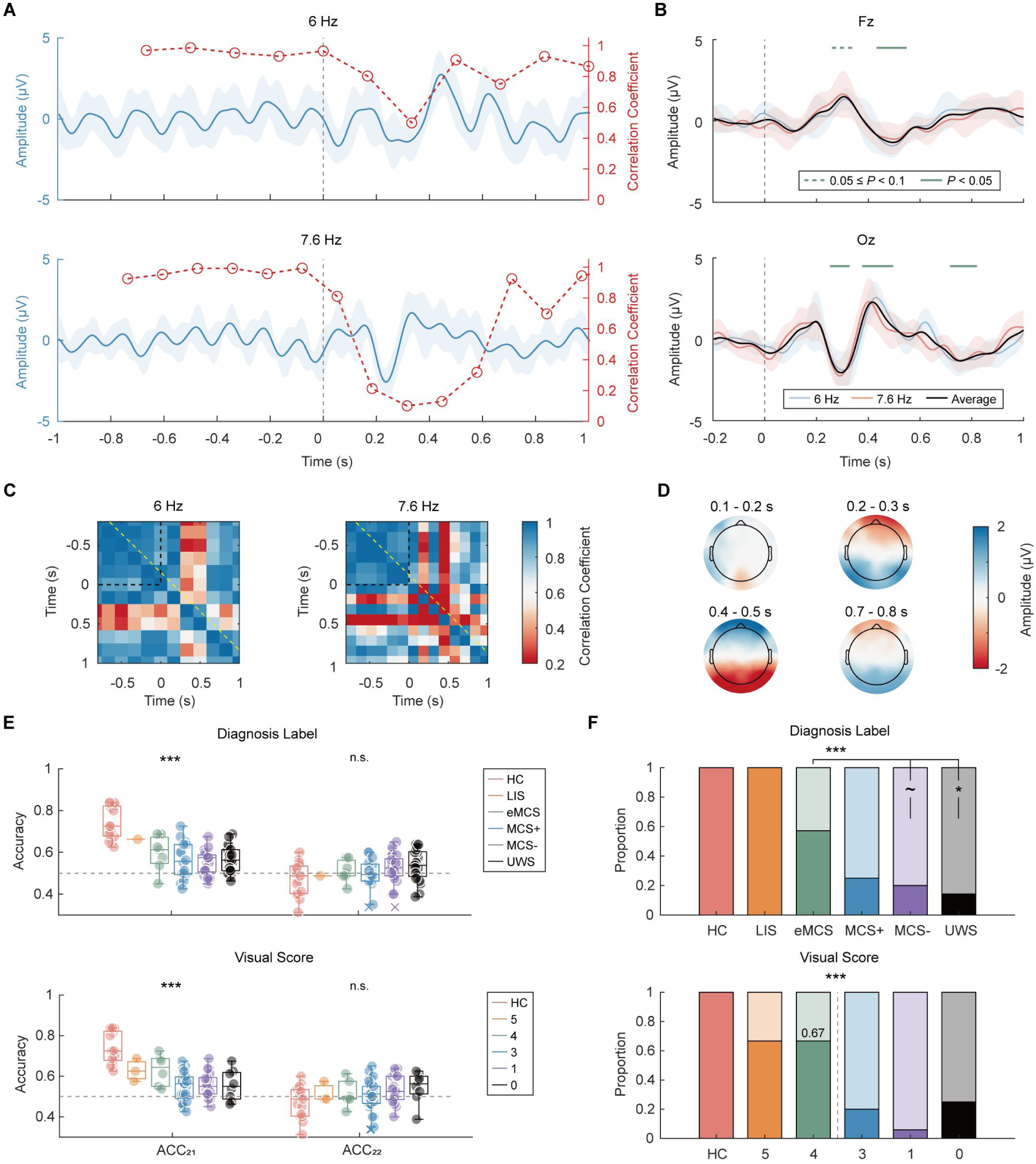
Analysis of the novel stimulus ERP and associated results across groups. In group HC, (**A**) waveforms and the inter-segment correlation curve at channel Oz before and after the onset of the novel stimulus; (**B**) ERP responses to the novel stimulus at channels Fz and Oz; (**C**) correlation matrix of different cycle segments at channel Oz (the yellow dashed line corresponds to the result of the red curve in panel **A**; (**D**) ERP topographic maps for corresponding time windows. For panels (**A**) and (**B**), shaded areas represent the standard error of the mean. (**E**) ACC_21_ and ACC_22_ values across different groups. Results of multiple comparisons (Bonferroni corrected) are shown in table S7. (**F**) The proportion of participants in each group who exhibit a significant ACC_21_ and a non-significant ACC_22_ (dark color) versus those who do not (light color). The grey dashed line in the lower panel demarcates groups with and without object discrimination behavior according to the CRS-R. Chi-squared test results: χ^2^ = 36.923, *P* < 0.001 for diagnostic labels (eMCS vs. MCS-: χ^2^ = 3.732, *P* = 0.053; eMCS vs. UWS: χ^2^ = 5.143, *P* = 0.023); for visual scores: χ^2^ = 40.877, *P* < 0.001. In panels (**E-F**): not significant (n.s.), *P* > 0.1; ∼, *P* < 0.1; *, *P* < 0.05; **, *P* < 0.01; ***, *P <* 0.001.

The analysis above confirmed that the novel stimulus elicited specific ERP activity. To objectively quantify visual discrimination in individual participants, we therefore employed a dual-classifier analysis comparing responses to the novel versus standard stimuli. Based on individual (Supplementary Fig. 2C) and group-averaged results (Supplementary Fig. 3B and C; Supplementary Table 6), the maximum ACC_21_ across the two stimulus conditions and the corresponding ACC_22_ were extracted for analysis. The results showed that ACC_21_ differed significantly across groups with different diagnosis levels [*F*(5, 79) = 16.195, *P* < 0.001, *η*^2^ = 0.506; one-way ANOVA] and visual scores [*F*(5, 78) = 19.267, *P* < 0.001, *η*^2^ = 0.553; one-way ANOVA], whereas ACC_22_ did not show significant differences across groups (Fig. 4E and Supplementary Table 7). Furthermore, the number of individuals with significant ACC_22_ and non-significant ACC_22_ also differed significantly across groups (for diagnostic labels: χ^2^ = 36.923, *P* < 0.001, Chi-squared test; for visual scores: χ^2^ = 40.877, *P* < 0.001, Chi-squared test; Fig. 4F). Such individuals constituted the majority in groups at the eMCS level and above and in those with a visual score greater than 4, but constituted only a small minority in the UWS, MCS groups, and in groups with a visual score less than 4. Specifically, the proportion in the eMCS group showed a (marginally) significant difference compared to the UWS (χ^2^ = 3.732, *P* = 0.053; Chi-squared test) and MCS- (χ^2^ = 5.143, *P* = 0.023; Chi-squared test) groups. These results indicate that ACC_21_ and ACC_22_ can further assess the state of consciousness, and their characterization of visual function aligns closely with the object recognition item of the CRS-R.

### Three-Level Integration: Visual Function, Consciousness State, and Prognosis Assessment

The p-value indicators corresponding to the three extracted levels were binarized to characterize whether a participant exhibited a significant response at each level (Fig. 5A): a value of 1 was assigned if the response was present, and 0 otherwise. Among the 85 participants, only 3 lacked a Level 1 response but possessed a Level 2 or 3 response; inspection revealed that these 3 participants (S14, S16, and S71) showed significant responses at the second harmonic for Level 1. Only 4 participants lacked a Level 2 response but possessed a Level 3 response (Supplementary Fig. 3E). Furthermore, the number of participants exhibiting a significant response decreased as the hierarchical level increased (Fig. 5A and Supplementary Fig. 3E). These results demonstrate that the visual functions indexed by our framework are hierarchical, consistent with the design intention.

**Figure 5.**
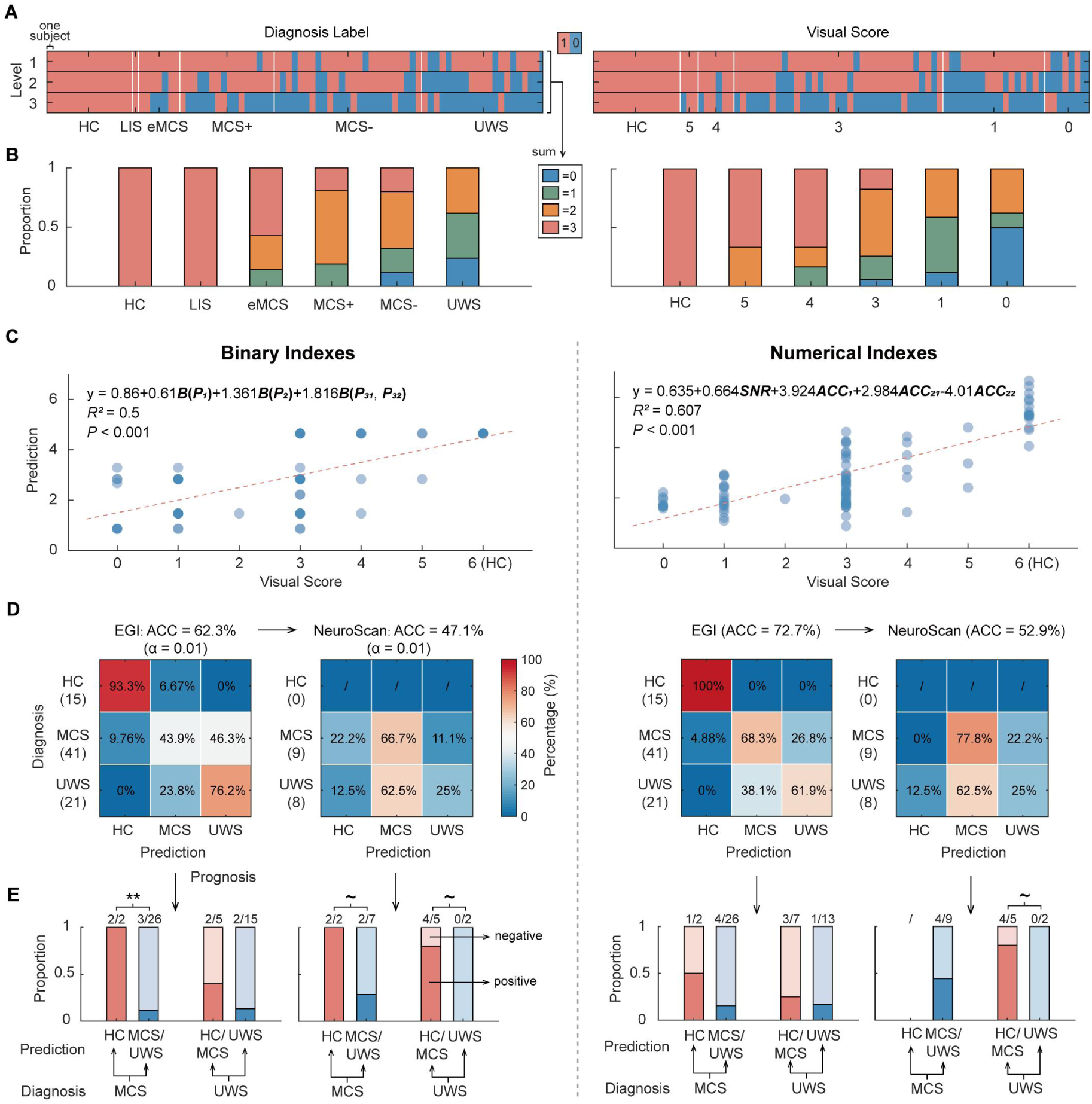
Comprehensive analysis of the three-level indexes. (**A**) Presence (coded as 1) or absence (coded as 0) of significant responses at each corresponding hierarchical level across different diagnostic and visual score groups. (**B**) Distribution of the number of hierarchical levels with significant responses across groups. (**C**) Regression analysis of binary and numerical indexes against visual scores. (**D**) Confusion matrices for three-class models based on binary indexes (left) and numerical indexes (right). (**E**) Proportion of individuals with favorable (dark shade) and unfavorable (light shade) prognosis, based on whether they were predicted by the models to have a higher consciousness state or the same/lower level. Binary-feature model, EGI cohort: MCS group (χ^2^ = 9.908, *P* = 0.002; Chi-squared test), UWS group (χ^2^ = 1.667, *P* = 0.197; Chi-squared test). Binary-feature model, Neuroscan cohort: MCS group (χ^2^ = 3.214, *P* = 0.073; Chi-squared test), UWS group (χ^2^ = 3.733, *P* = 0.053; Chi-squared test). Numerical-feature model, EGI cohort: MCS group (χ2 = 1.157, *P* = 0.218; Chi-squared test), UWS group (χ^2^ = 0.208, *P* = 0.648; Chi-squared test). Numerical-feature model, Neuroscan cohort: UWS group (χ^2^ = 3.733, *P* = 0.053; Chi-squared test). In panel (**E**): ∼, *P* < 0.1; *, *P* < 0.05; **, *P* < 0.01; ***, *P <* 0.001.

The binarized indicators from the three levels were summed to represent the number of hierarchical levels for which a participant showed a significant response. The results indicate that a higher diagnosis label or visual score is associated with a higher level of visual function (Fig. 5B). Across groups defined by diagnosis labels, all UWS patients failed to exhibit responses at all three levels simultaneously, whereas all HCs and the majority of eMCS patients showed concurrent responses at all three levels. MCS patients were predominantly characterized by significant responses at one or two levels.

Regression analyses between the visual scores and both the binarized indicators and the numerical metrics (SNR, ACC_1_, ACC_21_ and ACC_22_) showed that both types of indicators provided a good fit for the visual scores (Fig. 5C). The regression coefficients for Levels 2 and 3 were larger than that for Level 1, indicating a smaller contribution of Level 1 in signifying visual function. The coefficients for SNR, ACC_1_ and ACC_21_ were positive, contributing positively to the assessment of visual score, while the coefficient for ACC_22_ was negative, exerting a reverse effect—consistent with its role as a constraint metric.

To evaluate the efficacy of our paradigm in predicting the consciousness state, participants were divided into three groups: HC, MCS, and UWS. Two classification models were constructed based on the binarized indicators and the numerical metrics, respectively. The results showed that the accuracy of both models on the EGI data exceeded the chance level for a three-class task; however, the model based on binarized indicators underperformed the model based on numerical metrics (Fig. 5D). Both models demonstrated comparable performance when generalized to the data acquired with the Neuroscan system (Fig. 5D and Supplementary Fig. 1C).

Given that clinical behavioral scales are subject to rater subjectivity and patient motor function limitations, the diagnosis labels derived from these scales may not fully reflect the true state of consciousness. Therefore, we collected the clinical outcomes of DOC patients within three months post-experiment (Supplementary Table 1) and examined the concordance between the predictions of the different models and the prognosis. After excluding patients lost to follow-up, we found that patients predicted by the models to have a higher diagnosis level (e.g., UWS predicted as MCS) tended to have a better prognosis compared to those predicted to have the same or a lower diagnosis level (Fig. 5E). Notably, the priori knowledge-based model using binarized indicators identified more patients with good prognosis than the machine learning-based model using numerical metrics, in both the EGI and the generalized Neuroscan datasets.

## Discussion

This study proposes a hierarchical electrophysiological framework to evaluate visual function and consciousness in DOC patients. Utilizing a dual-target paradigm with statistical testing and machine learning, the framework assesses sensory input, selective attention, and visual discrimination. Results demonstrate that this hierarchical assessment shows high consistency with clinical behavioral scales while successfully detecting residual visual cognition and covert consciousness missed by the behavioral scale. Furthermore, patients categorized at higher diagnosis levels by the framework exhibited significantly better recovery trends, highlighting its prognostic value. Validated across different electrode types, this framework shows strong potential as an objective bedside tool.

### Neurocognitive Considerations of the Hierarchical Metrics

At the first level, our framework utilizes the SNR of the SSVEP to index basic sensory input function. The generation of SSVEPs depends on the integrity of the pathway from the retina to the primary visual cortex^37^ and is sensitive to impairment of the visual pathways ^53^. In this study, the vast majority of patients lacking a visual startle reflex failed to elicit significant SSVEP. This suggests potential damage to the visual pathways necessary for this activity, which may ultimately affect cortical function and structure^54^. A recent study also indicates the presence of varying degrees of visual pathway impairment in DOC patients^55^. Conversely, the ability to elicit significant SSVEP implies relative integrity of the primary sensory pathway, which forms the cornerstone for all higher-order visual cognition. The differences in VEP observed among DOC patients with varying degrees of injury^55–59^ likely reflect variations in the integrity of basic subcortical pathways and the excitability of the visual cortex, providing a preliminary physiological basis for assessing the state of consciousness.

At the second level, our framework assesses selective attention by decoding the location of the spatially attended target. The implementation of selective attention relies on a top-down control network, which includes the parietal cortex, prefrontal cortex, and the frontal eye field^60,61^. This network interacts with the visual cortex to enhance the neural representation of stimuli at the attended location^44^. In this study, we utilized the SSVEP as a signal carrier, capitalizing on its sensitivity to attentional allocation, to capture attentional activity. This includes large-angle gaze shifts, small-angle eye movements, and covert attention that occurs without eye movements. In our study, ACC_1_ demonstrated a strong association with the consciousness. This finding is not only consistent with previous reports of correlations between BCI accuracy (based on multiple visual targets) and consciousness^23–33^, but also aligns with the theoretical foundation that attention is a necessary condition for the emergence of consciousness^62^. The preservation of attentional function implies that visual input information is enhanced and maintained in cortical processing, which is a neural prerequisite for that information to potentially gain conscious representation^62^. Furthermore, a subset of patients exhibited significant ACC_1_ despite showing no observable visual tracking behavior, suggesting they retained covert attentional abilities typically missed by behavioral scales. In summary, the ACC_1_-related results in this study provide further electrophysiological evidence for understanding the relationship between consciousness and attention.

At the third level, our framework uses the accuracy (ACC_21_) of discriminating responses to novel versus standard stimuli to probe higher-level change detection and visual discrimination. The ERP components elicited by the novel stimulus have latencies greater than 100 ms, a time point shown in studies involving direct thalamic perturbation to be potentially related to the emergence of consciousness^63^. The early ERP components (∼100-250 ms) may include the visual mismatch negativity, reflecting the brain’s automatic detection of a change in the stimulus sequence. This process can be modulated by consciousness^64^ and involves occipital and prefrontal regions^49^, which is consistent with the findings of the present study. The later ERP components (∼300-600 ms) may include P3-family waves, associated with working memory updating and conscious perception, involving activations in frontal and parietal regions^65^. Previous studies reporting correlations of these components with consciousness in DOC have primarily been in the auditory modality^15–17,66–70^. Our study provides a complementary perspective from the visual modality. In this study, a small subset of patients with low diagnostic levels and visual scores still exhibited significant ACC_21_. This may reveal that they retain either automatic deviance detection or conscious visual discrimination, abilities difficult to capture with behavioral scales. We also found that a small number of patients showed significant Level 3 responses in the absence of a significant Level 2 response. Beyond potential limitations in oculomotor function, this finding prompts consideration of the hierarchical physiological relationship underlying visual selective attention and visual discrimination processes, providing a basis for further optimization of scale design. Moreover, to prevent false significance of ACC_21_ due to factors like eye closure, we used ACC_22_ as a constraint. The negative coefficient for this term in the regression analysis confirmed its effectiveness.

### Clinical Implications and Applications

This study provides objective electrophysiological evidence supporting the hierarchical design of the clinical CRS-R scale: the number of individuals with significant responses decreases as the hierarchical level increases, and those exhibiting significant responses at higher levels fundamentally possess significant responses at lower levels; furthermore, the proportion of participants with significant responses at each level largely aligns with the visual scores of the scale. Additionally, by utilizing EEG to overcome the scale’s reliance on (motor) behavioral performance, our paradigm discovered that a minority of DOC patients could “recognize objects” without showing “tracking” behavior, and that some patients with low behavioral consciousness states also retained higher-level functions. These findings suggest that these patients may retain covert consciousness, but their motor impairments preclude its behavioral expression. This CMD condition^12^ is not detectable by the CRS-R scale. Compared to recent studies of a similar nature, our study achieved a relatively high detection rate of potential consciousness (individuals with significant responses at more than one level) within a relatively large sample size (Supplementary Table 8), further underscoring its applicability in the clinical assessment of DOC.

In terms of consciousness classification, the three-class machine learning model based on the three-level numerical metrics achieved an accuracy of 70%, comparable to a previous study^21^. Moreover, this model achieved above-chance accuracy when generalized to the cross-device data, indicating that it possesses a certain capability for consciousness prediction. The model based on binary indicators underperformed the model based on numerical metrics, especially when generalized to data with lower impedance and higher stability, suggesting that the predetermined confidence level captures only part of the data features. However, the advantage of the binary indicator model is that it allows for rapid individual assessment without requiring time-consuming training, making it more suitable for preliminary evaluation of admitted patients in clinical settings or for large-scale community screening.

More importantly, regardless of the model, the DOC group predicted to have a higher consciousness state contained a larger proportion of patients with good prognosis, indicating that patients classified as having better consciousness by the models tend to have a better prognosis. This finding is consistent with prior evidence that CMD patients who could operate a visual BCI had better prognostic outcomes^13^. In prognosis assessment, compared to the numerical metric model, the binary indicator model appeared to predict a greater number of patients with good prognosis. This suggests, on one hand, that the machine learning model based on behavioral labels may be affected by label inaccuracy, whereas the model based on prior knowledge might better reflect the true condition of DOC patients. On the other hand, it demonstrates the good clinical applicability of our framework and its potential utility in addressing challenges in DOC diagnosis and prognosis evaluation.

The feature extraction and model establishment in this study were all based on data from nine occipital channels. Reducing or increasing the number of channels (Supplementary Fig. 1) had almost no effect on the three-level metrics at the group level, but it did affect the values for individual participants (Supplementary Fig. 4A-C). The magnitude of the channel selection effect may depend on the relative position between the patient’s gaze point and the stimulus target^71^. Incomplete channel coverage may fail to capture the stimulus projection area on the scalp, which leaves room for improvement in the stimulus presentation and channel selection algorithms. In terms of model classification and prognosis assessment, changes in the number of channels had little impact on the overall performance and generalizability of the models (Supplementary Fig. 4D and E). This indicates that the framework have the potential to be adapted to EEG systems with different channel layouts, facilitating adoption across hospitals. Furthermore, the correlations of age, onset duration, and etiology with the metrics at each level were weak (Supplementary Fig. 5), suggesting the framework’s suitability for bedside examination of DOC patients with varying physical conditions. Finally, by assessing three levels of visual function within a single unified paradigm, our framework saves valuable time for both healthcare providers and patients compared to previous approaches requiring multiple separate tests.

### Limitations

The participants enrolled in this study were all patients capable of eye-opening, which may limit the application scenarios of this paradigm. Put differently, this paradigm might be used to explore the functional performance of patients unable to open their eyes voluntarily under passive eye-opening conditions. Additionally, the sample sizes for some DOC subgroups (e.g., eMCS) were relatively small, which may be insufficient to reflect the true situation of these categories. It is important to note that not all consciousness or visual features detected by the clinical behavioral scale were identified by our paradigm. This discrepancy may stem, on one hand, from fluctuations in the consciousness state of DOC patients, which repeated measurements might mitigate^72^. On the other hand, EEG is susceptible to various non-pathological factors; therefore, non-significant response results should be interpreted with caution^10^, and combining other detection methods could further improve assessment accuracy.

## Data availability

Because of ethical considerations, the human data are protected and available under restricted access according to the data security guidelines of Beijing Tiantan Hospital. Data access can be obtained by contacting upon reasonable request.

## Acknowledgements

We thank Peijian Sun, Shuai Han, Xiaojun Lin, Xueling Chen, and Xiaoli Geng from Beijing Tiantan Hospital for their assistance in the clinical data collection for this study. We also thank Bowen Li, Changxing Huang, Yonghao Song, and Xiaoyang Li from Tsinghua University, and Nanlin Shi from Duke University for their suggestions on data processing and analysis.

