## Supplementary for "A Hierarchical Visual EEG Framework for the Assessment of Disorders of Consciousness"

Yuzhen Chen *et al.*

Corresponding author:

Steven Laureys,

Xiaogang Chen,

Jianghong He,

Xiaorong Gao,

### **The file includes:**

Supplementary Methods: Evoked Potential Identification

Supplementary Figures 1 to 6

Supplementary Tables 1 to 8

### METHODS

#### Evoked Potential Identification

The paradigm incorporates three stimulus elements corresponding to the three hierarchical levels of visual function: sensory input, selective attention, and visual discrimination. The evoked potentials associated with each level possess distinct characteristics and require different methods for detection.

***1) Sensory Input***. The two RSVP targets presented at fixed frequencies induce a frequency-following effect in the visual system related to sensory input. Therefore, the presence of a SSVEP response at the corresponding frequency in the neural activity of visual regions can be used to determine whether a participant possesses basic sensory input function. This SSVEP response can be characterized by its SNR at the target frequency. To mitigate potential variability from individual electrodes, we employed CCA[41] to integrate information from nine occipital channels (O1, O2, Oz, POz, Pz, PO7, PO3, PO4, PO6). Specifically, the data from these nine channels form the signal matrix $X$. The other set of variables consists of reference signals $Y_i$ corresponding to the stimulation frequency $f_i$, defined using sine-cosine templates at that frequency:

$$Y_i = \begin{bmatrix} sin(2\pi f_i t) \\ cos(2\pi f_i t) \\ \vdots \\ sin(2\pi N_h f_i t) \\ cos(2\pi N_h f_i t) \end{bmatrix} \quad (1)$$

where $f_i$ is the stimulation frequency, and $N_h$ is the number of harmonics. $N_h$ was set to 1 for constructing the CCA coefficient spectrum. CCA finds the weight vectors $w_x$ and $w_y$ for $X$ and $Y$, respectively, to maximize the correlation coefficient between their linear combinations $x = X^T w_x$ and $y = Y^T w_y$:

$$\rho(x, y) = \max_{w_x, w_y} \frac{E[w_x^T X Y^T w_y^T]}{\sqrt{E[w_x^T X X^T w_x^T] E[w_y^T Y Y^T w_y^T]}} \quad (2)$$

Here, $\rho$ is the maximum canonical correlation coefficient between variables $X$ and $Y$. A CCA coefficient spectrum is constructed by calculating the canonical correlation coefficient between $X$ and reference signals corresponding to a range of frequencies (0.2, 0.4, 0.6, ..., 39.8, 40.0 Hz). The CCA SNR is then defined in a manner analogous to the traditional SNR[39], specifically as the ratio of the CCA coefficient at the target frequency to the mean CCA coefficient of the surrounding frequencies[40]:

$$CCA\,SNR = \frac{M \cdot C(f_0)}{\sum_{m}^{M/2} [(C(f_0) - \Delta f \cdot m) + (C(f_0) + \Delta f \cdot m)]} \tag{3}$$

where $f_0$ is the specified stimulation frequency, $\Delta f$ is the frequency resolution of the CCA spectrum (set to 0.2 Hz), $C(f)$ denotes the CCA coefficient at a specific frequency, and $M$ is set to 10, corresponding to frequencies within ±1 Hz around the target frequency.

Considering the potentially limited eye movement capability and visual field in some DOC patients, SSVEP responses to both stimulation frequencies (f1, f2) under both conditions (colored pictures on the left or right) are included in the analysis. This yields four CCA SNR (hereafter referred to as SNR) metrics: left-f1, left-f2, right-f1, right-f2. For each trial, these four metrics are calculated to form four SNR sequences. The sequence with the strongest average response is selected. A one-tailed t-test is performed against the null response level (H1: SNR > 1) for this sequence to obtain the p-value for the SSVEP response[42], denoted as $P_1$. A confidence level of $\alpha = 0.05$ is used as the criterion for determining a significant individual SSVEP response. If a participant's $P_1$ value is less than α, the participant is considered to possess basic visual sensory input function.

***2) Selective Attention***: The paradigm uses two distinct frequencies to tag the left and right targets, eliciting differentiable SSVEP responses containing two frequency components. Recognition algorithms commonly used in SSVEP-BCI can thus be employed to determine whether a participant can attend to the instructed target, thereby assessing their visual selective attention or visual tracking ability. To compare the performance of different algorithm types, we employed one training-free algorithm (FBCCA[45]) and two training-based algorithms (TRCA[46] and TDCA[47]) commonly used in the SSVEP-BCI field. The specific algorithmic principles are as follows:

- FBCCA[45]: FBCCA is an extension of CCA. Based on CCA, the signal matrix $X$ is decomposed into multiple sub-band signals via a filter bank. For the n-th sub-band signal $X_n$, its CCA coefficient with the reference signal $Y_i$ in equation (1) is calculated. Thus, for the i-th stimulation frequency $f_i$, the correlation coefficient vector consists of n coefficients:

$$\rho_i = \begin{bmatrix} \rho_i^1 \\ \vdots \\ \rho_i^n \end{bmatrix} = \begin{bmatrix} \rho(X_1^T w_x(X_1 Y_i), Y^T w_y(X_1 Y_i)) \\ \vdots \\ \rho(X_N^T w_x(X_{N_b} Y_i), Y^T w_y(X_{N_b} Y_i)) \end{bmatrix} \tag{4}$$

where $\rho(x, y)$ denotes the correlation coefficient between $x$ and $y$; $i$ = 1, 2 correspond to the left and right targets (i.e., $f_1$ = 6 and $f_2$ = 7.6); $N_b$ is the number of sub-bands, which, along with the number of harmonics $N_h$ in equation (1), was set to 4. A weighted sum of this coefficient vector serves as the final feature for target identification, with the weighting scheme following a previous report[45].

- TRCA[46]: Signals in evoked potentials are considered a superposition of task-related signal components and task-unrelated noise components. Due to the phase-locking property of evoked potentials, the signal components exhibit high consistency across trials. The idea of TRCA is to extract the evoked potential component by maximizing the trial-to-trial consistency of the signal $S$. For each class, TRCA aims to maximize the inter-trial covariance:

$$w^T S w = \sum_{\substack{h_1,h_2=1 \\ h_1 \neq h_2}}^{N_t} \sum_{j_1,j_2=1}^{N_{ch}} w_{j_1} w_{j_2} Cov\left(x_{j_1}^{(h_1)}(t), x_{j_2}^{(h_2)}(t)\right) \tag{5}$$

where $h$ denotes the h-th trial, $N_t$ is the total number of trials; $j$ denotes the j-th channel, and $N_{ch}$ is the total number of channels. This optimization problem is solved by incorporating a constraint on finite variance:

$$w^T Q w = \sum_{j_1,j_2=1}^{N_{ch}} w_{j_1} w_{j_2} Cov\left(x_{j_1}(t), x_{j_2}(t)\right) \tag{6}$$

The spatial filter (channel weights) $w$ is thus obtained by:

$$\hat{w} = \underset{w}{arg\,max}\frac{w^T S w}{w^T Q w} \tag{7}$$

Applying the above procedure to training data yields a spatial filter and a corresponding signal template for each class (different stimulation frequency). Applying the spatial filter to test data and calculating its correlation with the templates provides the final feature for target identification. This study employed the standard TRCA version without ensemble strategies.

- TDCA[47]: SSVEP responses to different frequencies may share a common spatial pattern. Based on this, TDCA aims to find a common spatial filter $w^T$ across classes that minimizes the within-class variance while maximizing the between-class variance for the multi-class visual responses. Specifically, the between-class matrix and within-class matrix are computed as:

$$H_b = \frac{1}{\sqrt{N_c}}[\bar{X}^1 - \bar{X}^{all}, \cdots, \bar{X}^{N_c} - \bar{X}^{all}] \tag{8}$$

$$H_w = \frac{1}{\sqrt{N_t}}[X^{(1)} - \bar{X}^{(1)}, \cdots, X^{(N_t)} - \bar{X}^{(N_t)}] \tag{9}$$

where $X^i$ denotes the data for the i-th class, $X^{all}$ denotes the data for all classes, and $X^{(h)}$ denotes the data for the h-th trial; $N_c$ is the number of classes, and $N_t$ is the number of trials. According to the Fisher criterion, the spatial filter $W$ can be obtained by optimizing the following objective:

$$\underset{W}{maximize}\frac{tr(W^T S_b W)}{tr(W^T S_w W)} \tag{10}$$

where $S_b$ and $S_w$ are the between-class and within-class covariance matrices, respectively:

$$S_b = H_b H_b^T \tag{11}$$

$$S_w = H_w H_w^T \tag{12}$$

Applying the above procedure to training data yields a common spatial filter for all classes. Applying this filter to the averaged data of each class from the training set yields the signal template for that class. Applying the common spatial filter to test data and calculating its correlation with the templates provides the final feature for target identification. This study employed the standard TDCA version without data augmentation strategies[47].

Each block in the paradigm includes stimulation for two classes: attending to the left target and attending to the right target. Therefore, the algorithm's recognition accuracy in the experiment indicates whether the participant can select the target as instructed. The training-free algorithm directly calculates an accuracy for each block's data. The training-based algorithms calculate accuracy for each block treated as test data using a leave-one-block-out cross-validation strategy. Since the accuracy generally shows an increasing trend with longer attentional time, the best average accuracy from the final second across the three algorithms is used as the metric, denoted as $ACC_1$. The sequence of $ACC_1$ values from all blocks is obtained. Following statistical testing approaches from previous SSVEP work[13], a one-tailed t-test is performed on the $ACC_1$ sequence against the chance level for a binary task (H1: ACC > 0.5), yielding a corresponding p-value, denoted as $P_2$. A confidence level of $\alpha = 0.05$ is used as the criterion for determining a significant individual $ACC_1$. If a participant's $P_2$ value is less than $\alpha$, the participant is considered to possess visual selective attention ability.

***3) Visual Discrimination***: The colored abstract picture sequence (standard stimulus) used in this paradigm contains an embedded concrete human face picture (novel stimulus). This design can induce an SSVEP in the visual system, as well as a transient event-related potential (ERP) related to the function of visual discrimination. To determine whether the ERP corresponding to the novel stimulus is significant at the individual level, the following procedure was implemented:

First, three time points were selected, and a 1-second epoch of data following each was extracted: the onset of an abstract picture presented before the face picture, the onset of the face picture itself, and the onset of an abstract picture presented after the face picture. These two abstract pictures were chosen to ensure their onsets were separated from the face picture onset by at least 1 second. These three epochs were labeled as: *standard (pre)*, *novelty*, and *standard (post)*.

Subsequently, the TRCA algorithm was employed to train two classifiers: one to discriminate between *standard (pre)* and *novelty* epochs, and another to discriminate between *standard* *(pre)* and *standard (post)* epochs. Using a leave-one-out cross-validation strategy, two sequences of accuracy values were obtained, denoted as $ACC_{21}$ and $ACC_{22}$, respectively.

Considering the potentially limited eye movement capability and visual field in some DOC patients, for the analysis, we selected the $ACC_{21}$ sequence with the best average accuracy from the two conditions (target on left and target on right), along with the $ACC_{22}$ sequence from the corresponding condition, as the metrics.

One-tailed t-tests were performed on the $ACC_{21}$ and $ACC_{22}$ sequences against the chance level for a binary classification task (H1: ACC > 0.5), yielding two corresponding p-values, denoted as $P_{31}$ and $P_{32}$. A confidence level of $\alpha = 0.05$ was used as the criterion for determining whether an individual's $ACC_{21}$ and $ACC_{22}$ were significant: If a participant's $P_{31}$ value was less than α and his $P_{32}$ value was greater than α, the participant was considered to possess visual discrimination ability; If a participant's $P_{31}$ and $P_{32}$ values were both greater than α, it suggested that the observed difference between the *standard (pre)* and *novelty* data might be due to other factors (e.g., gaze shift, eye closure) rather than a genuine neural response to the face picture.

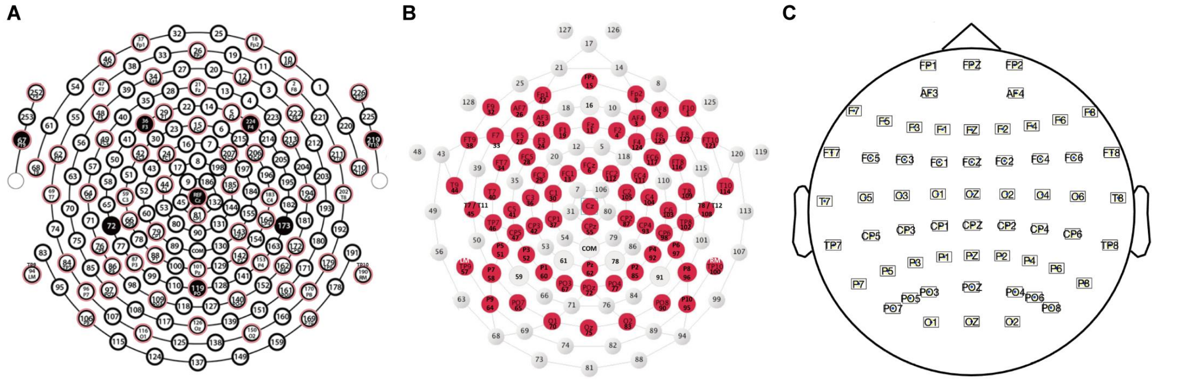


**Supplementary Figure 1 Three EEG channel configurations.** (**A**) The 256-channel configuration of the EGI system (180 channels remain after removing cheek channels). The channels used for feature extraction and for analyzing the effect of channel selection are: O1, O2, Oz, POz (E116, E150, E126, E119); Pz, PO7, PO3, PO4, PO6 (E101, E97, E109, E140, E161); E108, E117, E139, E151. (**B**) The 128-channel configuration of the EGI system. The channels used for feature extraction and for analyzing the effect of channel selection are: O1, O2, Oz, POz (E70, E83, E75, E72); Pz, PO7, PO3, PO4, PO6 (E62, E65, E67, E77, E90); E66, E71, E76, E84. (**C**) The 64-channel configuration of the Neuroscan system. The channels used for feature extraction are: O1, O2, Oz, POz, Pz, PO7, PO3, PO4, PO6.

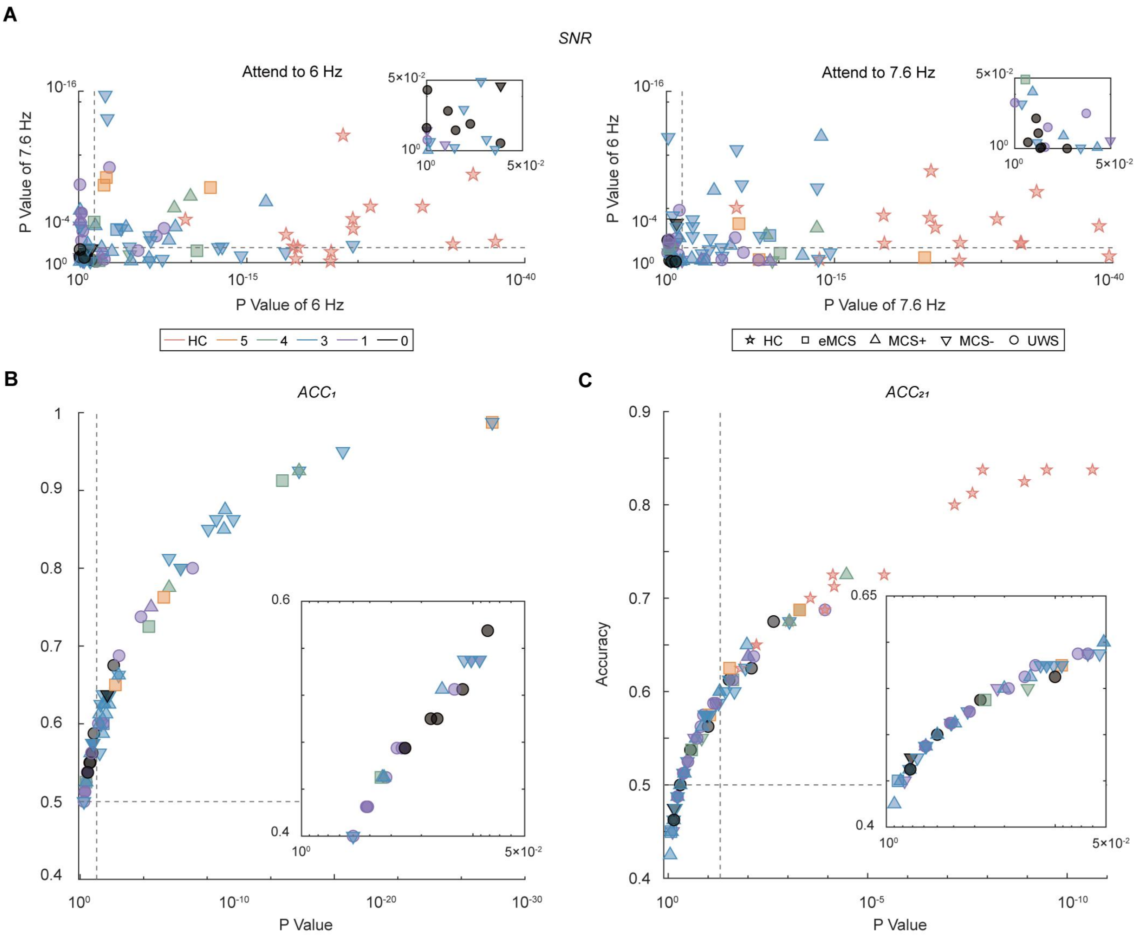


**Supplementary Figure 2 Individual results of the three-level metrics.** (**A**) Significance p-values of the SNR for the two stimulation frequencies, under conditions where the colored pictures appeared on the left (6 Hz) or right (7.6 Hz) side. (**B**) Accuracy ($ACC_1$) of the classifier discriminating responses to left versus right attended targets and its significance. (**C**) Accuracy ($ACC_{21}$) of the classifier discriminating responses to the novel versus standard stimuli and its significance. Insets in all panels show magnified views of the corresponding results.

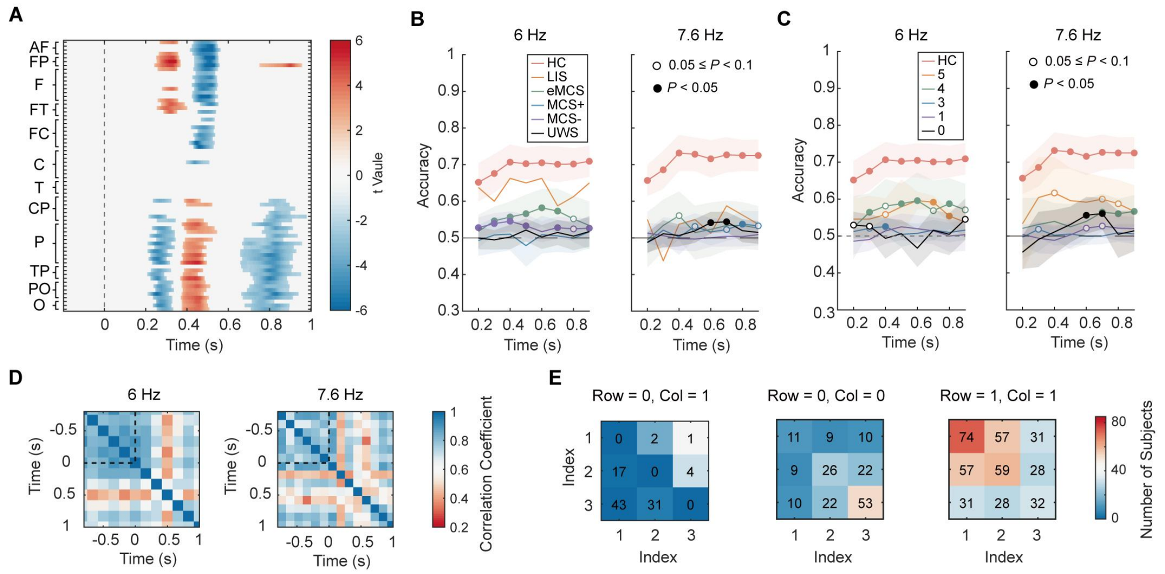


**Supplementary Figure 3 Responses to the novel stimulus and participant counts under specific criteria.** (**A**) Statistical test results (significance map) of the ERP response to the novel stimulus across all channels in group HC. Only areas with $P < 0.05$ are displayed. Temporal dynamics of ACC21 when attending to targets on different sides for (**B**) different diagnosis label groups and (**C**), different visual score groups. In panels (**B**, **C**), responses with $P < 0.05$, $P < 0.01$, and $P < 0.001$ are all indicated by a solid circle (•). (**D**), Correlation matrix of different cycle segments, averaged across all channels, for HCs. (**E**) Distribution of participant counts with significant or non-significant responses at each hierarchical level. '1' and '0' denote the presence and absence of a significant response at that level, respectively.

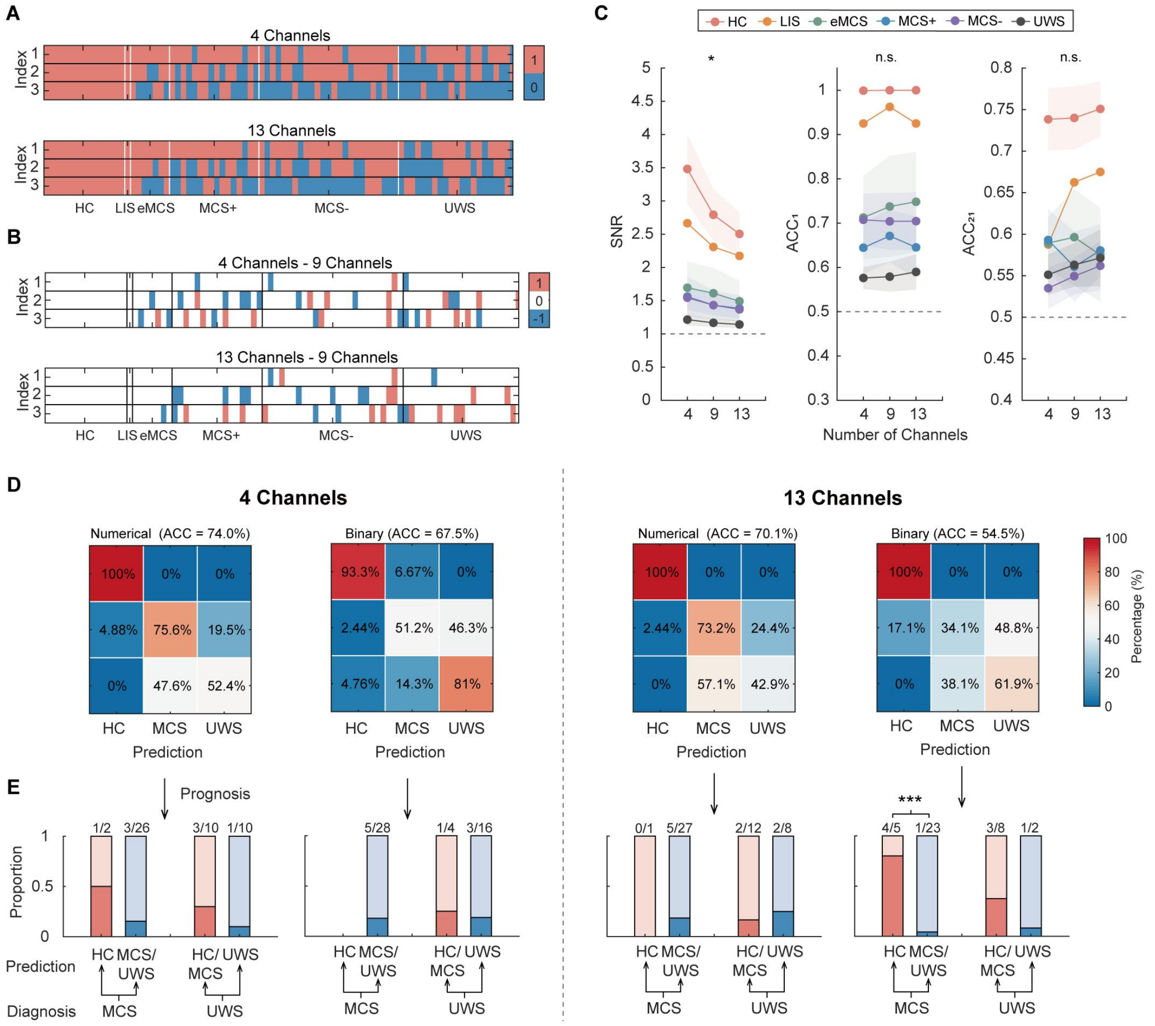


**Supplementary Figure 4 Influence of channel selection on the results.** (**A**) Metrics for each hierarchical level extracted using either 4 or 13 occipital channels (Supplementary Fig. 1) and (**B**) their comparison with the results obtained using 9 channels. (**C**) Changes in the numerical values of the hierarchical-level metrics under different numbers of selected channels. (**D**) Model performance and (**E**) corresponding prognosis statistics for models using 4 or 13 occipital channels. For the numerical-feature model: In the 4-channel cohort, $\chi^2$ = 1.157, $P$ = 0.218 for the MCS group and $\chi^2$ = 1.250, $P$ = 0.264 for the UWS group; In the 13-channel cohort, $\chi^2$ = 0.635, $P$ = 0.225 for the MCS group and $\chi^2$ = 0.648, $P$ = 0.208 for the UWS group. For the binary-feature model: In the 4-channel cohort, $\chi^2$ = 0.078, $P$ = 0.780 for the UWS group; In the 13-channel cohort, $\chi^2$ = 16.025, $P$ < 0.001 for the MCS group and $\chi^2$ = 2.552, $P$ = 0.110 for the UWS group. In panel (**E**): ~, $P$ < 0.1; *, $P$ < 0.05; **, $P$ < 0.01; ***, $P$ < 0.001.

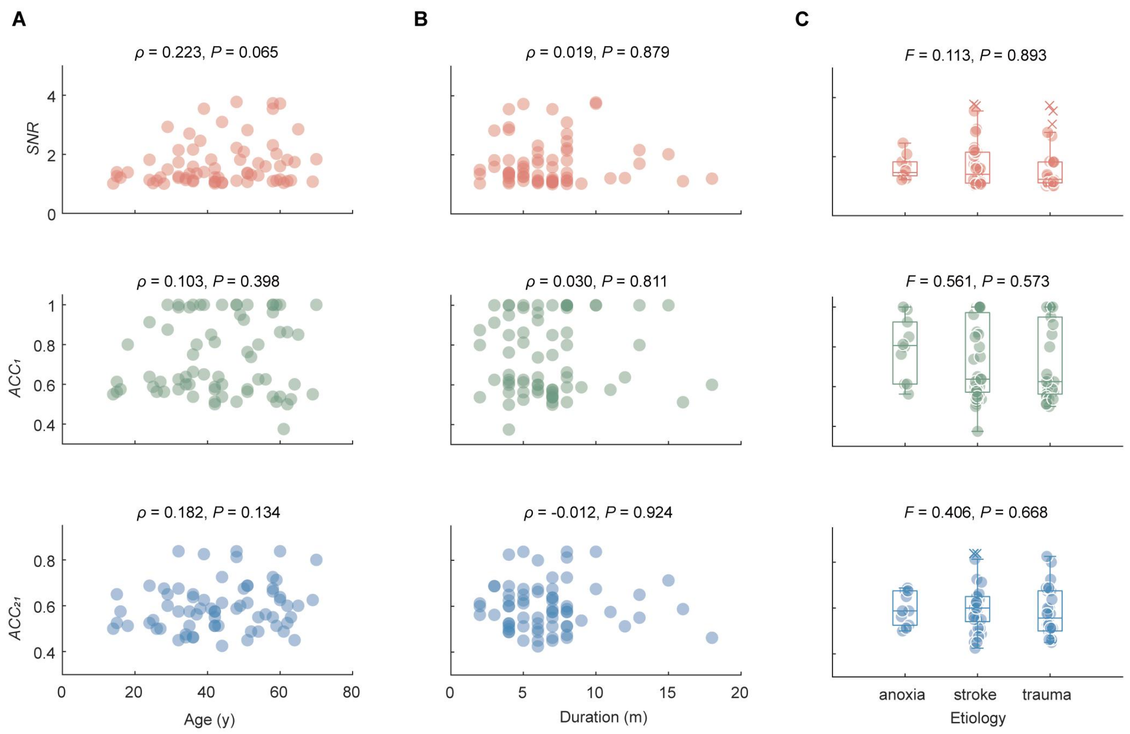


**Supplementary Figure 5 Relationship between different factors and the hierarchical-level metrics.** The three factors analyzed are: (**A**) age, (**B**) onset duration, and (**C**) etiology of the DOC patients.

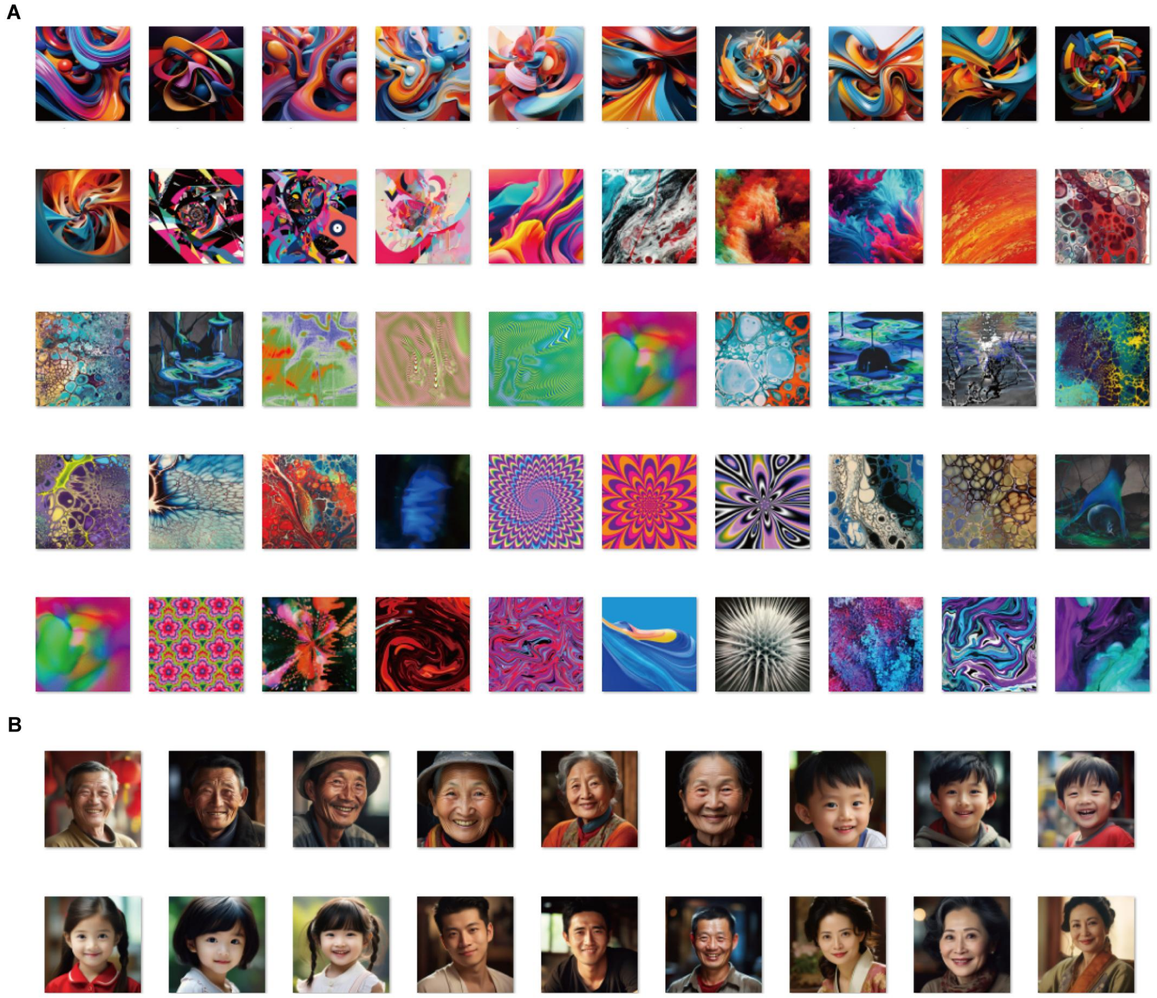

**Supplementary Figure 6 Stimulus materials.** (**A**) Abstract color pictures used as the standard stimulus. (**B**) Face color pictures used as the novel stimulus. Facial images shown are computer-generated (via *Midjourney*) and are not photographs of real people.

**Supplementary Table 1 Summary of the Subject Information**

| Subject | Gender | Age | Diagnosis | CRS-R Score | Etiology | Duration(m) | Prognosis | Channel Number |
|---|---|---|---|---|---|---|---|---|
| S1 | f | 41–45 | MCS- | 032102 | stroke | 8 | negative | 256 |
| S2 | m | 51–55 | MCS- | 132102 | trauma | 3 | positive | 256 |
| S3 | m | 26–30 | HC | / | / | / | / | 256 |
| S4 | f | 51–55 | MCS- | 245103 | trauma | 4 | negative | 256 |
| S5 | m | 41–45 | MCS- | 132202 | stroke | 12 | negative | 256 |
| S6 | m | 56–60 | MCS- | 031102 | stroke | 6 | negative | 256 |
| S7 | m | 26–30 | HC | / | / | / | / | 128 |
| S8 | f | 56–60 | MCS- | 103102 | stroke | 5 | negative | 128 |
| S9 | m | 31–35 | HC | / | / | / | / | 128 |
| S10 | m | 31–35 | HC | / | / | / | / | 256 |
| S11 | f | 56–60 | UWS | 012102 | trauma | 7 | negative | 128 |
| S12 | m | 70–74 | UWS | 002001 | trauma | 6 | negative | 128 |
| S13 | m | 35–39 | MCS+ | 332102 | stroke | 13 | unknown | 128 |
| S14 | m | 30–34 | MCS+ | 434102 | trauma | 5 | negative | 128 |
| S15 | m | 35–39 | MCS+ | 334102 | stroke | 4 | negative | 256 |
| S16 | m | 15–19 | UWS | 102102 | trauma | 8 | negative | 128 |
| S17 | m | 40–44 | UWS | 112102 | trauma | 8 | negative | 256 |
| S18 | m | 30–34 | HC | / | / | / | / | 256 |
| S19 | m | 30–34 | eMCS | 454123 | stroke | 11 | / | 256 |
| S20 | m | 20–24 | MCS+ | 136102 | trauma | 6 | negative | 256 |
| S21 | f | 60–64 | eMCS | 446112 | trauma | 8 | / | 256 |
| S22 | m | 40–44 | MCS- | 031102 | stroke | 7 | positive | 256 |
| S23 | f | 30–34 | MCS+ | 432102 | anoxia | 5 | unknown | 256 |
| S24 | f | 60–64 | MCS+ | 115101 | stroke | 4 | positive | 256 |
| S25 | m | 50–54 | MCS- | 032101 | stroke | 5 | negative | 256 |
| S26 | m | 55–59 | MCS- | 132101 | stroke | 7 | unknown | 256 |
| S27 | m | 50–54 | LIS | 002101 | stroke | 4 | / | 256 |
| S28 | m | 25–29 | MCS- | 132101 | stroke | 2 | unknown | 256 |
| S29 | m | 50–54 | eMCS | 452122 | stroke | 6 | / | 256 |
| S30 | f | 50–54 | HC | / | / | / | / | 256 |
| S31 | m | 25–29 | HC | / | / | / | / | 256 |
| S32 | m | 35–39 | MCS+ | 332102 | stroke | 7 | negative | 256 |
| S33 | f | 30–34 | HC | / | / | / | / | 256 |
| S34 | m | 30–34 | eMCS | 344212 | trauma | 18 | / | 256 |
| S35 | f | 30–34 | UWS | 002102 | stroke | 8 | negative | 256 |
| S36 | m | 25–29 | MCS+ | 335313 | trauma | 6 | unknown | 256 |
| S37 | m | 35–39 | MCS- | 133202 | trauma | 8 | negative | 256 |
| S38 | m | 25–29 | MCS+ | 332202 | trauma | 9 | unknown | 256 |
| S39 | m | 40–44 | MCS+ | 345113 | stroke | 6 | unknown | 256 |
| S40 | m | 35–39 | MCS+ | 332112 | anoxia | 8 | unknown | 256 |
| S41 | m | 50–54 | UWS | 002102 | anoxia | 6 | negative | 256 |
| S42 | f | 40–44 | MCS- | 232102 | stroke | 6 | negative | 256 |
| S43 | f | 65–69 | UWS | 112102 | stroke | 7 | negative | 256 |
| S44 | m | 55–59 | UWS | 112102 | stroke | 3 | unknown | 256 |
| S45 | m | 35–39 | MCS- | 235112 | anoxia | 2 | unknown | 256 |
| S46 | m | 35–39 | MCS- | 132102 | trauma | 7 | negative | 256 |
| S47 | f | 15–19 | UWS | 102102 | anoxia | 5 | negative | 256 |

**Supplementary Table 1** (Continued)

| Subject | Sex | Age | Diagnosis | CRS-R Score | Etiology | Duration(m) | Prognosis | Channel Number |
|---|---|---|---|---|---|---|---|---|
| S48 | f | 70–74 | UWS | 112102 | anoxia | 4 | negative | 256 |
| S49 | m | 35–39 | MCS+ | 332112 | stroke | 16 | negative | 256 |
| S50 | m | 30–34 | eMCS | 456212 | trauma | 7 | / | 256 |
| S51 | m | 35–39 | UWS | 112102 | anoxia | 4 | negative | 256 |
| S52 | m | 15–19 | MCS+ | 444213 | stroke | 2 | unknown | 256 |
| S53 | m | 40–44 | MCS- | 113102 | trauma | 8 | unknown | 256 |
| S54 | f | 30–34 | MCS- | 134003 | stroke | 7 | unknown | 256 |
| S55 | m | 30–34 | MCS+ | 334102 | trauma | 7 | negative | 256 |
| S56 | f | 20–24 | MCS- | 234102 | trauma | 4 | negative | 256 |
| S57 | m | 60–64 | MCS- | 233103 | anoxia | 4 | unknown | 256 |
| S58 | m | 40–44 | HC | / | / | / | unknown | 256 |
| S59 | m | 30–34 | eMCS | 236102 | stroke | 10 | / | 256 |
| S60 | m | 60–64 | MCS- | 232102 | anoxia | 7 | negative | 256 |
| S61 | f | 50–54 | UWS | 112102 | stroke | 7 | positive | 256 |
| S62 | m | 55–59 | UWS | 002102 | trauma | 7 | negative | 256 |
| S63 | m | 50–54 | HC | / | / | / | / | 256 |
| S64 | m | 25–29 | HC | / | / | / | / | 256 |
| S65 | m | 50–54 | HC | / | / | / | / | 256 |
| S66 | m | 50–54 | HC | / | / | / | / | 256 |
| S67 | m | 25–29 | UWS | 122102 | stroke | 8 | negative | 256 |
| S68 | m | 35–39 | UWS | 212102 | trauma | 4 | positive | 256 |
| S69 | m | 30–34 | MCS+ | 333102 | trauma | 4 | negative | 256 |
| S70 | m | 30–34 | UWS | 212102 | trauma | 4 | positive | 256 |
| S71 | m | 30–34 | MCS- | 133102 | stroke | 13 | negative | 256 |
| S72 | m | 25–29 | HC | / | / | / | / | 256 |
| S73 | m | 35–39 | HC | / | / | / | / | 256 |
| S74 | m | 25–29 | MCS- | 132102 | stroke | 4 | positive | 256 |
| S75 | m | 40–44 | MCS+ | 334102 | stroke | 6 | negative | 256 |
| S76 | f | 35–39 | eMCS | 443322 | stroke | 5 | / | 256 |
| S77 | f | 50–54 | UWS | 112102 | trauma | 3 | positive | 256 |
| S78 | m | 40–44 | UWS | 102102 | stroke | 4 | negative | 256 |
| S79 | f | 65–69 | MCS- | 232102 | trauma | 8 | negative | 256 |
| S80 | m | 55–59 | UWS | 102102 | stroke | 15 | negative | 256 |
| S81 | m | 35–39 | MCS- | 132102 | trauma | 40 | negative | 256 |
| S82 | f | 35–39 | MCS- | 133202 | trauma | 10 | positive | 256 |
| S83 | m | 15–19 | UWS | 112102 | stroke | 8 | negative | 256 |
| S84 | m | 25–29 | UWS | 111102 | anoxia | 5 | negative | 256 |
| S85 | m | 45–49 | MCS- | 114102 | stroke | 5 | negative | 256 |
| S86 | m | 15–19 | MCS- | 032102 | stroke | 6 | negative | 64 |
| S87 | m | 50–54 | MCS- | 132102 | stroke | 4 | positive | 64 |
| S88 | m | 40–44 | UWS | 012102 | stroke | 5 | negative | 64 |
| S89 | f | 35–39 | MCS- | 032102 | stroke | 6 | negative | 64 |
| S90 | m | 55–59 | UWS | 002102 | stroke | 4 | positive | 64 |
| S91 | f | 25–29 | UWS | 102102 | anoxia | 5 | positive | 64 |
| S92 | m | 60–64 | MCS- | 132102 | anoxia | 5 | negative | 64 |
| S93 | m | 15–19 | MCS- | 232102 | trauma | 5 | negative | 64 |
| S94 | m | 30–34 | MCS- | 032102 | stroke | 4 | positive | 64 |

**Supplementary Table 1** (Continued)

| Subject | Sex | Age | Diagnosis | CRS-R Score | Etiology | Duration(m) | Prognosis | Channel Number |
|---|---|---|---|---|---|---|---|---|
| S95 | f | 60–64 | UWS | 102100 | stroke | 3 | positive | 64 |
| S96 | m | 30–34 | MCS- | 123102 | trauma | 16 | positive | 64 |
| S97 | m | 55–59 | MCS- | 234102 | stroke | 5 | positive | 64 |
| S98 | f | 45–49 | UWS | 112002 | anoxia | 7 | negative | 64 |
| S99 | m | 40–44 | UWS | 112102 | stroke | 11 | unknown | 64 |
| S100 | f | 45–49 | MCS- | 132102 | stroke | 7 | negative | 64 |
| S101 | f | 50–54 | UWS | 112012 | trauma | 19 | positive | 64 |
| S102 | f | 30–34 | UWS | 112102 | trauma | 7 | negative | 64 |

### Supplementary Table 2 Statistical Test Results of p-Values for SNR in Different Groups

| Harmonic Multiplier | Diagnosis Label | | | | | | | | | | | |
|---|---|---|---|---|---|---|---|---|---|---|---|---|
| | HC | | LIS | | eMCS | | MCS+ | | MCS- | | UWS | |
| | f1 | f2 | f1 | f2 | f1 | f2 | f1 | f2 | f1 | f2 | f1 | f2 |
| ×1 | 0.000 | 0.000 | / | / | 0.028 | 0.029 | 0.008 | 0.002 | 0.001 | 0.000 | 0.037 | 0.017 |
| ×2 | 0.000 | 0.000 | / | / | 0.025 | 0.004 | 0.006 | 0.001 | 0.000 | 0.001 | 0.261 | 0.134 |
| ×3 | 0.000 | 0.000 | / | / | 0.091 | 0.019 | 0.002 | 0.003 | 0.001 | 0.032 | 0.351 | 0.076 |
| ×4 | 0.000 | 0.000 | / | / | 0.099 | 0.005 | 0.027 | 0.166 | 0.001 | 0.059 | 0.097 | 0.188 |

| Harmonic Multiplier | Visual Score | | | | | | | | | |
|---|---|---|---|---|---|---|---|---|---|---|
| | 5 | | 4 | | 3 | | 1(/2) | | 0 | |
| | f1 | f2 | f1 | f2 | f1 | f2 | f1 | f2 | f1 | f2 |
| ×1 | 0.140 | 0.114 | 0.013 | 0.020 | 0.000 | 0.000 | 0.049 | 0.007 | 0.462 | 0.343 |
| ×2 | 0.188 | 0.039 | 0.010 | 0.012 | 0.000 | 0.000 | 0.139 | 0.061 | 0.678 | 0.360 |
| ×3 | 0.127 | 0.051 | 0.019 | 0.044 | 0.000 | 0.014 | 0.178 | 0.062 | 0.169 | 0.018 |
| ×4 | 0.061 | 0.064 | 0.070 | 0.047 | 0.001 | 0.051 | 0.050 | 0.102 | 0.177 | 0.334 |

Values of 0.000 correspond to $P < 0.001$.

### Supplementary Table 3 Statistical Test Results for SNR Across Different Groups

| Multiple Comparison (Bonferroni corrected) | | | | | |
|---|---|---|---|---|---|
| Diagnosis Label | | | Visual Score | | |
| **Group 1** | **Group 2** | **p** | **Group 1** | **Group 2** | **p** |
| HC | LIS | 1.000 | HC | 5 | 0.026 |
| HC | eMCS | 0.000 | HC | 4 | 0.000 |
| HC | MCS+ | 0.000 | HC | 3 | 0.000 |
| HC | MCS- | 0.000 | HC | 1 | 0.000 |
| HC | UWS | 0.000 | HC | 0 | 0.000 |
| LIS | eMCS | 1.000 | 5 | 4 | 1.000 |
| LIS | MCS+ | 1.000 | 5 | 3 | 1.000 |
| LIS | MCS- | 1.000 | 5 | 1 | 0.692 |
| LIS | UWS | 0.329 | 5 | 0 | 0.273 |
| eMCS | MCS+ | 1.000 | 4 | 3 | 1.000 |
| eMCS | MCS- | 1.000 | 4 | 1 | 1.000 |
| eMCS | UWS | 0.535 | 4 | 0 | 0.649 |
| MCS+ | MCS- | 1.000 | 3 | 1 | 1.000 |
| MCS+ | UWS | 1.000 | 3 | 0 | 0.661 |
| MCS- | UWS | 0.955 | 1 | 0 | 1.000 |

Values of 0.000 correspond to $P < 0.001$.

### Supplementary Table 4 Statistical Test Results for $ACC_1$ in Different Groups

| Time (s) | Diagnosis Label | | | | | | Visual Score | | | | |
|---|---|---|---|---|---|---|---|---|---|---|---|
| | HC | LIS | eMCS | MCS+ | MCS- | UWS | 5 | 4 | 3 | 1 | 0 |
| FBCCA | | | | | | | | | | | |
| 1 | 0.000 | / | 0.383 | 0.283 | 0.937 | 0.936 | 0.280 | 0.517 | 0.709 | 0.928 | 0.941 |
| 2 | 0.000 | / | 0.416 | 0.089 | 0.661 | 0.944 | 0.478 | 0.112 | 0.462 | 0.919 | 0.925 |
| 3 | 0.000 | / | 0.182 | 0.047 | 0.667 | 0.980 | 0.377 | 0.036 | 0.389 | 0.978 | 0.867 |
| 4 | 0.000 | / | 0.319 | 0.079 | 0.533 | 0.999 | 0.492 | 0.046 | 0.310 | 0.997 | 0.992 |
| 5 | 0.000 | / | 0.325 | 0.030 | 0.438 | 0.996 | 0.368 | 0.137 | 0.198 | 0.994 | 0.911 |
| TRCA | | | | | | | | | | | |
| 1 | 0.000 | / | 0.178 | 0.012 | 0.000 | 0.274 | 0.050 | 0.215 | 0.000 | 0.071 | 0.667 |
| 2 | 0.000 | / | 0.032 | 0.010 | 0.000 | 0.486 | 0.075 | 0.029 | 0.000 | 0.135 | 0.500 |
| 3 | 0.000 | / | 0.060 | 0.009 | 0.000 | 0.134 | 0.059 | 0.037 | 0.000 | 0.158 | 0.194 |
| 4 | 0.000 | / | 0.028 | 0.020 | 0.000 | 0.033 | 0.099 | 0.013 | 0.000 | 0.001 | 0.424 |
| 5 | 0.000 | / | 0.008 | 0.014 | 0.000 | 0.034 | 0.056 | 0.003 | 0.000 | 0.003 | 0.344 |
| TDCA | | | | | | | | | | | |
| 1 | 0.000 | / | 0.083 | 0.015 | 0.001 | 0.240 | 0.000 | 0.149 | 0.036 | 0.001 | 0.013 |
| 2 | 0.000 | / | 0.202 | 0.006 | 0.000 | 0.270 | 0.000 | 0.100 | 0.156 | 0.000 | 0.235 |
| 3 | 0.000 | / | 0.053 | 0.005 | 0.000 | 0.165 | 0.000 | 0.072 | 0.024 | 0.000 | 0.069 |
| 4 | 0.000 | / | 0.038 | 0.021 | 0.000 | 0.232 | 0.000 | 0.089 | 0.011 | 0.000 | 0.114 |
| 5 | 0.000 | / | 0.006 | 0.009 | 0.000 | 0.347 | 0.000 | 0.047 | 0.007 | 0.000 | 0.133 |

Values of 0.000 correspond to $P < 0.001$.

**Supplementary Table 5 Statistical Test Results for $ACC_1$ Across Different Groups**

| Multiple Comparison (Bonferroni corrected) | | | | | |
|---|---|---|---|---|---|
| Diagnosis Label | | | Visual Score | | |
| **Group 1** | **Group 2** | **p** | **Group 1** | **Group 2** | **p** |
| HC | LIS | 1.000 | HC | 5 | 0.159 |
| HC | eMCS | 0.000 | HC | 4 | 0.004 |
| HC | MCS+ | 0.000 | HC | 3 | 0.000 |
| HC | MCS- | 0.000 | HC | 1 | 0.000 |
| HC | UWS | 0.000 | HC | 0 | 0.000 |
| LIS | eMCS | 1.000 | 5 | 4 | 1.000 |
| LIS | MCS+ | 0.365 | 5 | 3 | 1.000 |
| LIS | MCS- | 0.643 | 5 | 1 | 0.111 |
| LIS | UWS | 0.048 | 5 | 0 | 0.130 |
| eMCS | MCS+ | 1.000 | 4 | 3 | 1.000 |
| eMCS | MCS- | 1.000 | 4 | 1 | 0.027 |
| eMCS | UWS | 0.064 | 4 | 0 | 0.050 |
| MCS+ | MCS- | 1.000 | 3 | 1 | 0.193 |
| MCS+ | UWS | 0.409 | 3 | 0 | 0.485 |
| MCS- | UWS | 0.015 | 1 | 0 | 1.000 |

Values of 0.000 correspond to $P < 0.001$.

### Supplementary Table 6 Statistical Test Results for $ACC_{21}$ in Different Groups

| Time (s) | Diagnosis Label | | | | | | | | | | | |
|---|---|---|---|---|---|---|---|---|---|---|---|---|
| | Attend to f1 | | | | | | Attend to f2 | | | | | |
| | HC | LIS | eMCS | MCS+ | MCS- | UWS | HC | LIS | eMCS | MCS+ | MCS- | UWS |
| 0.2 | 0.000 | 1.000 | 0.250 | 0.624 | 0.031 | 0.515 | 0.000 | 1.000 | 0.549 | 0.750 | 0.178 | 0.804 |
| 0.3 | 0.000 | 1.000 | 0.036 | 0.381 | 0.008 | 0.648 | 0.000 | 1.000 | 0.207 | 0.134 | 0.363 | 0.282 |
| 0.4 | 0.000 | 1.000 | 0.024 | 0.333 | 0.002 | 0.446 | 0.000 | 1.000 | 0.073 | 0.464 | 0.547 | 0.339 |
| 0.5 | 0.000 | 1.000 | 0.030 | 0.754 | 0.028 | 0.101 | 0.000 | 1.000 | 0.368 | 0.060 | 0.567 | 0.137 |
| 0.6 | 0.000 | 1.000 | 0.013 | 0.377 | 0.132 | 0.445 | 0.000 | 1.000 | 0.293 | 0.209 | 0.485 | 0.007 |
| 0.7 | 0.000 | 1.000 | 0.037 | 0.407 | 0.045 | 0.207 | 0.000 | 1.000 | 0.198 | 0.088 | 0.379 | 0.003 |
| 0.8 | 0.000 | 1.000 | 0.092 | 0.514 | 0.069 | 0.408 | 0.000 | 1.000 | 0.186 | 0.035 | 0.401 | 0.121 |
| 0.9 | 0.000 | 1.000 | 0.192 | 0.248 | 0.038 | 0.215 | 0.000 | 1.000 | 0.224 | 0.072 | 0.283 | 0.183 |
| Time (s) | Visual Score | | | | | | | | | | | |
| | | 5 | 4 | 3 | 1 | 0 | | 5 | 4 | 3 | 1 | 0 |
| 0.2 | | 0.156 | 0.324 | 0.166 | 0.771 | 0.086 | | 0.359 | 0.334 | 0.615 | 0.378 | 0.941 |
| 0.3 | | 0.184 | 0.037 | 0.085 | 0.724 | 0.087 | | 0.133 | 0.238 | 0.053 | 0.606 | 0.588 |
| 0.4 | | 0.083 | 0.052 | 0.027 | 0.227 | 0.580 | | 0.097 | 0.106 | 0.441 | 0.723 | 0.373 |
| 0.5 | | 0.115 | 0.017 | 0.396 | 0.102 | 0.384 | | 0.112 | 0.249 | 0.359 | 0.437 | 0.137 |
| 0.6 | | 0.012 | 0.037 | 0.301 | 0.157 | 0.862 | | 0.102 | 0.148 | 0.512 | 0.097 | 0.038 |
| 0.7 | | 0.007 | 0.096 | 0.132 | 0.238 | 0.310 | | 0.060 | 0.035 | 0.500 | 0.059 | 0.017 |
| 0.8 | | 0.020 | 0.036 | 0.242 | 0.500 | 0.441 | | 0.081 | 0.022 | 0.234 | 0.120 | 0.448 |
| 0.9 | | 0.162 | 0.066 | 0.128 | 0.438 | 0.090 | | 0.135 | 0.021 | 0.342 | 0.126 | 0.325 |

Values of 0.000 correspond to $P < 0.001$.

### Supplementary Table 7 Statistical Test Results for $ACC_{21}$ across Different Groups

| Multiple Comparison (Bonferroni corrected) | | | | | |
|---|---|---|---|---|---|
| Diagnosis Label | | | Visual Score | | |
| Group 1 | Group 2 | p | Group 1 | Group 2 | p |
| HC | LIS | 1.000 | HC | 5 | 0.179 |
| HC | eMCS | 0.001 | HC | 4 | 0.021 |
| HC | MCS+ | 0.000 | HC | 3 | 0.000 |
| HC | MCS- | 0.000 | HC | 1 | 0.000 |
| HC | UWS | 0.000 | HC | 0 | 0.000 |
| LIS | eMCS | 1.000 | 5 | 4 | 1.000 |
| LIS | MCS+ | 1.000 | 5 | 3 | 0.675 |
| LIS | MCS- | 1.000 | 5 | 1 | 1.000 |
| LIS | UWS | 1.000 | 5 | 0 | 1.000 |
| eMCS | MCS+ | 1.000 | 4 | 3 | 0.086 |
| eMCS | MCS- | 1.000 | 4 | 1 | 0.396 |
| eMCS | UWS | 1.000 | 4 | 0 | 0.672 |
| MCS+ | MCS- | 1.000 | 3 | 1 | 1.000 |
| MCS+ | UWS | 1.000 | 3 | 0 | 1.000 |
| MCS- | UWS | 1.000 | 1 | 0 | 1.000 |

Values of 0.000 correspond to $P < 0.001$.

**Supplementary Table 8 Proportion of Patients with Potential Consciousness across Diagnostic Groups in Various Studies**

| | eMCS | MCS | UWS | Visual Score > 2 |
|---|---|---|---|---|
| Pan et al., 2014 | / | 3/3 (100%) | 1/4 (25%) | 3/3 (100%) |
| Li et al., 2015 | 2/2 (100%) | 1/3 (33.3%) | 2/6 (33.3%) | 3/4 (75%) |
| Xiao et al., 2018 | 0/1 (0%) | 2/5 (40%) | 1/8 (12.5%) | 2/4 (50%) |
| Xiao et al., 2018 | 1/1 (100%) | 5/6 (83.3%) | 4/6 (66.7%) | 2/4 (50%) |
| Xie et al., 2018 | / | 2/3 (66.7%) | 1/5 (25%) | 3/4 (75%) |
| Pan et al., 2020 [1] | / | 7/18 (38.9%) | 11/30 (36.7%) | 6/13 (46.2%) |
| Huang et al., 2021 | / | 3/4 (75%) | 0/3 (0%) | 2/2 (100%) |
| Xiao et al., 2022 | / | 6/8 (75%) | 5/10 (50%) | 5/7 (71.4%) |
| Pan et al., 2023a | / | 2/4 (50%) | 1/8 (12.5%) | / |
| Pan et al., 2023b | / | 2/5 (40%) | 1/3 (33.3%) | 2/3 (66.7%) |
| Yi et al., 2024 [2] | / | 2/8 (25%) | 0/1 (0%) | 2/8 (25%) |
| **this article** | **6/7 (85.7%)** | **30/41 (73.1%)** | **8/21 (38.1%)** | **34/44 (77.3%)** |

[1] Visual paradigm only. [2] EEG only.
